# Population impact of new TB vaccines may depend on efficacy against infectious asymptomatic TB: a modelling study

**DOI:** 10.1101/2025.03.24.25323816

**Authors:** Hira Tanvir, Rebecca A Clark, Tom Sumner, Katherine C Horton, Tomos O Prŷs-Jones, Roel Bakker, Kirankumar Rade, Vidya Mave, Mark Hatherill, Gavin Churchyard, Rein MGJ Houben, Richard G White

**Affiliations:** TB Modelling Group, TB Centre, and Centre for Mathematical Modelling of Infectious Diseases, Department of Infectious Disease Epidemiology, London School of Hygiene & Tropical Medicine, London, UK; Vaccine Centre, London School of Hygiene & Tropical Medicine, London, UK; KNCV Tuberculosis Foundation, The Hague, Netherlands; Independent Consultant, New Delhi, India; Byramjee–Jeejeebhoy Government Medical College, Johns Hopkins University Clinical Research Site, Pune, India; South African Tuberculosis Vaccine Initiative, Institute of Infectious Disease and Molecular Medicine and Department of Pathology, University of Cape Town, Cape Town, South Africa; Aurum Institute NPC, Houghton, Parktown, South Africa; School of Public Health, University of Witwatersrand, Johannesburg, South Africa; Department of Medicine, Vanderbilt University, Nashville, Tennessee, USA

**Author notes:** Co-primary authors (Hira Tanvir, Rebecca A Clark).

## Abstract

Tuberculosis (TB) remains a leading cause of infectious disease death. Modelling predicts new TB vaccines may reduce global burden but rely on assumptions about vaccine efficacy by TB disease stage and TB natural history, which may be incorrect. We explored the sensitivity of estimates of the impact of new TB vaccines to uncertainties in efficacy by disease stage and natural history.

We developed a dynamic compartmental TB model for India, including early TB disease stages. Scenarios assumed 50% vaccine efficacy for 10 years and prevented progression to a) only infectious symptomatic disease, or b) any infectious disease, or c) any disease. We estimated impact on averting disease episodes over 2030–2050, compared to no-new-vaccine introduction.

Results suggest, over three years, there was little difference in the proportion of cumulative symptomatic disease episodes averted by vaccines preventing only infectious symptomatic disease, any infectious disease, or any disease (1.8%, 2.3%, and 2.4%, respectively). However, over 20 years, compared to vaccines preventing only infectious symptomatic disease, vaccines preventing any infectious disease, or any disease, averted a markedly higher proportion of symptomatic disease episodes (8.2%, 21.0%, and 25.1%, respectively), due to preventing continued transmission from infectious asymptomatic disease.

The population impact of new TB vaccines may depend on efficacy against infectious asymptomatic disease. TB vaccine trials should measure impact on infectious asymptomatic disease to enable better estimates of the potential value of new TB vaccines. Further data collection is required to better understand the transmissibility, morbidity, and dynamics of asymptomatic disease.

## Background

Tuberculosis (TB) causes around 10.8 million new episodes and 1.25 million deaths annually (1). The highest number of TB episodes and deaths occurs in India, with more than a quarter of global TB incidence and mortality (1). There is evidence that infectious asymptomatic TB disease contributes to transmission and may be a significant driver of the TB epidemic (2,3). Data from national TB prevalence surveys suggests that infectious asymptomatic TB may account for at least 50% of prevalent bacteriologically positive disease (4). Asymptomatic TB may also progress to symptomatic disease and contribute to post TB morbidity (3–7).

New TB vaccines may be available soon, with a recently completed phase 2b trial of the vaccine candidate M72/AS01_E_ demonstrating encouraging results of around 50% (95% confidence interval = 2.1%–74.2%) impact on the incidence of symptomatic disease (8). The primary endpoint of this trial was efficacy against bacteriologically confirmed (infectious) symptomatic TB (previously referred to as *clinical TB*), but the impact on infectious asymptomatic TB was not measured (5,9,10). The phase 3 trial is ongoing (11).

Recent modelling studies suggest that introducing a new vaccine that prevents TB could yield substantial global health and economic benefits (12–16). However, these estimates did not incorporate current uncertainty in the natural history of asymptomatic TB or uncertainty in the efficacy of the vaccine by TB disease stage (12–16). If new TB vaccines have no impact on progression to non-infectious and/or infectious asymptomatic TB, and/or infectious asymptomatic TB contributes to transmission differently than assumed, these estimates of the impact of new TB vaccines could be incorrect (5). Incorporating uncertainty in the infectiousness of asymptomatic TB and the impact of the vaccine by disease stages is thus critical for understanding the potential impact of new TB vaccines.

We explored the sensitivity of estimates of the impact of new TB vaccines to uncertainties in vaccine efficacy by TB disease stage and infectiousness of asymptomatic TB with India as a case study, as its high TB burden, capacity to develop and test vaccines, and the likelihood of early adoption of new TB vaccines, make India a suitable population for this research question.

## Methods

### Setting and empirical data

We used demographic projections for India by single age and year from the United Nations population data (17). Estimates for TB incidence, TB case notifications, and TB mortality were from the World Health Organization (WHO) (18). TB disease and infection prevalence estimates were from the India National TB Prevalence Survey and Pandey et al, 2017 (19,20). A full list of parameters and sources is in Supplementary Table S 1.

### Model structure

We used an age-stratified compartmental transmission model of TB, updating the natural history in line with recent modelling (Figure 1a) (12,21). Our natural history structure consisted of nine compartments, modelling pathways from uninfected (S), to *Mtb* infection (I, from which self-clearance of *Mtb* infection to a cleared compartment [C] was possible), through disease stages (described below), treatment (T), and recovery (R and Rt). See supporting materials for full details.

**Figure 1.**
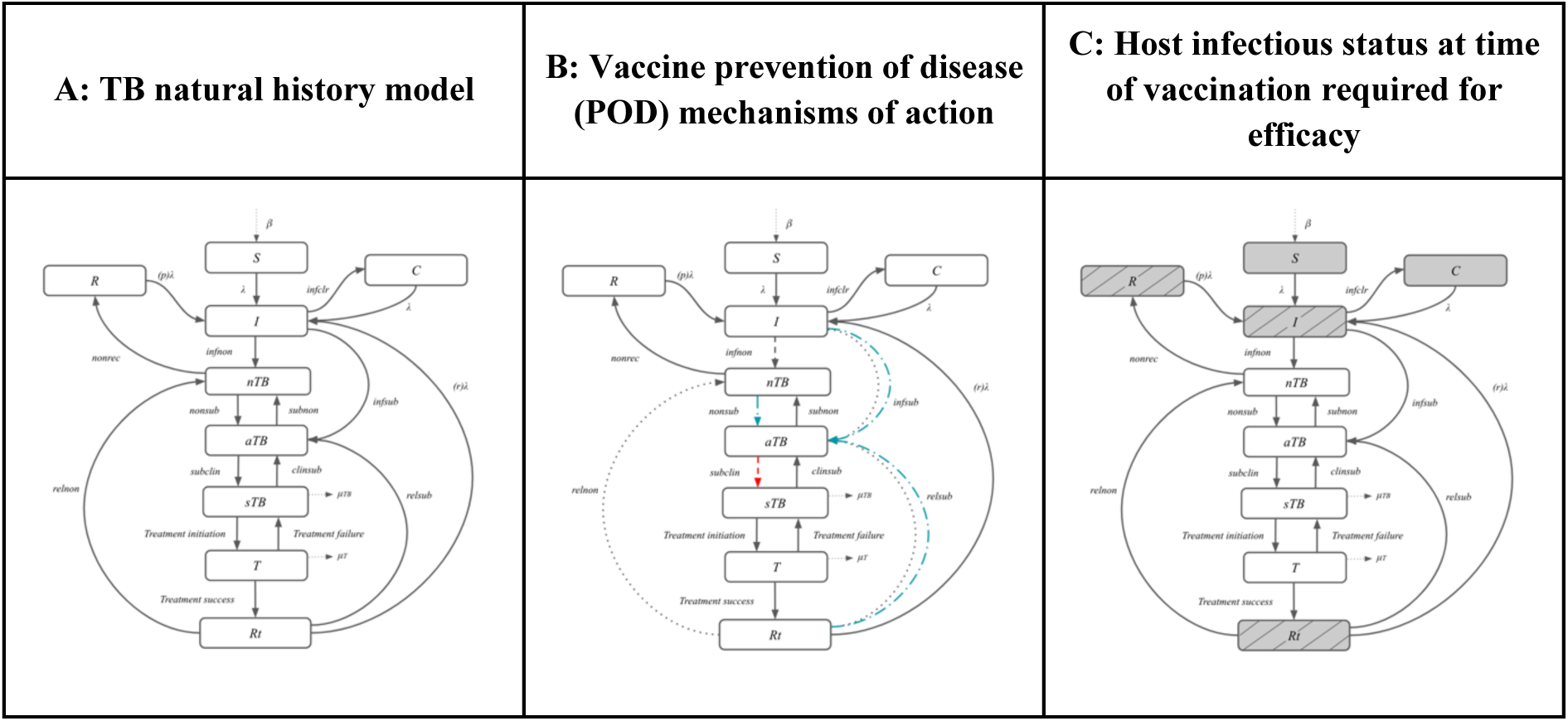
**a)** Illustration of core TB natural history model **(b)** Scenarios for prevention of disease: preventing infectious symptomatic disease only (red dashed line), preventing any infectious disease (blue dash-dotted lines) or preventing any disease (grey dotted lines). **(c)** Illustration of host infectious status at time of vaccination required for efficacy – Default: any infection status (AI, all grey shaded boxes) or sensitivity analysis: current infection status only (CI, only grey shaded boxes with lines). *Abbreviations: S = Susceptible; I = Infection; C = Cleared; R = Recovered; nTB = Non-infectious Disease; aTB = Infectious Asymptomatic Disease; sTB = Infectious Symptomatic Disease; T = On-Treatment; Rt = Recovered after treatment. Dashed lines indicate the mechanism of action of each vaccine type: dashed red lines show the transition impacted for a vaccine preventing progression to symptomatic disease, blue dash-dotted blue lines for a vaccine preventing any infectious disease, and grey dotted lines for a vaccine preventing any disease stage. Shaded boxes indicate the host infectious status at time of vaccination required for efficacy*.

In line with emerging evidence and recent WHO guidance, we represented TB disease with three stages: infectious symptomatic TB (sTB), infectious asymptomatic TB (aTB), and non-infectious TB (nTB) (9,10). We defined individuals with sTB as individuals with bacteriologically positive (infectious) TB reporting symptoms suggestive of TB during screening. We assumed additional TB mortality, regression to aTB, and treatment initiation from sTB. We defined aTB as bacteriologically positive (infectious) TB in individuals not reporting symptoms suggestive of TB during screening. To be conservative, we assumed no additional mortality from aTB, and we assumed the relative infectiousness of aTB compared to sTB was between 0.61 and 1 (2). We assumed further disease progression from aTB to sTB, and regression from aTB to nTB. In our model, we defined nTB as bacteriologically negative (non-infectious) disease, which could be asymptomatic or symptomatic.

We allowed for recovery from nTB, and further disease progression to aTB. To be conservative, we assumed no additional mortality from nTB.

### Calibration

The model was calibrated using history matching with emulation (22) using the *hmer* R package (23) to obtain 500 parameter sets fitted to all 14 calibration targets (Table S 5). Targets for all ages were the TB incidence rate in 2000 and 2020, mortality rate in 2000 and 2020, case notification rate in 2000 and 2020, infectious TB prevalence in 2015 and 2021, and infectious asymptomatic-to-symptomatic TB prevalence ratio in 2020 (4,17,19,20,24–27). Targets for children were the incidence rate and case notification rate in 2020. Targets for adults were the TB incidence rate and case notification rate in 2020 and active tuberculosis prevalence rate in 2021.

### No-new-vaccine scenario

We simulated a *no-new-vaccine* scenario from 1900–2050 assuming the constraints above and that the quality and coverage of non-vaccine TB interventions post 2022 would be maintained at 2022 levels, and no new TB vaccine introduction.

### New TB vaccine policy scenarios

Using the calibrated *no-new-vaccine scenario* model, we projected the impact of *Basecase* scenarios of the introduction of a new vaccine between 2030 and 2050, with vaccine characteristics informed by the M72/AS01_E_ phase IIb trial (8) and expert opinion (28) (Table 1). We assumed all vaccines had 50% efficacy to prevent disease (using different disease definitions as described below) if given to infected or uninfected individuals at the time of vaccination but not to individuals with nTB, aTB, or sTB (i.e., an ‘any infection’ vaccine (12)), and the duration of protection was 10 years on average with exponential waning (8,29). To illustrate the potential epidemiologic impact, we assumed a supply and resource unconstrained maximal rollout of the vaccines. We assumed all vaccines would be delivered routinely to those aged 15 (reaching 80% coverage linearly over 5 years) starting in 2030, and as two campaigns for ages 16–44 (reaching 70% coverage linearly over 5 years) in 2030 and 2040. See Supplementary Material section 4.2 for full details. Trends in the proportion of the population vaccinated over time are shown in Supplementary Figure S 7.

**Table 1.**
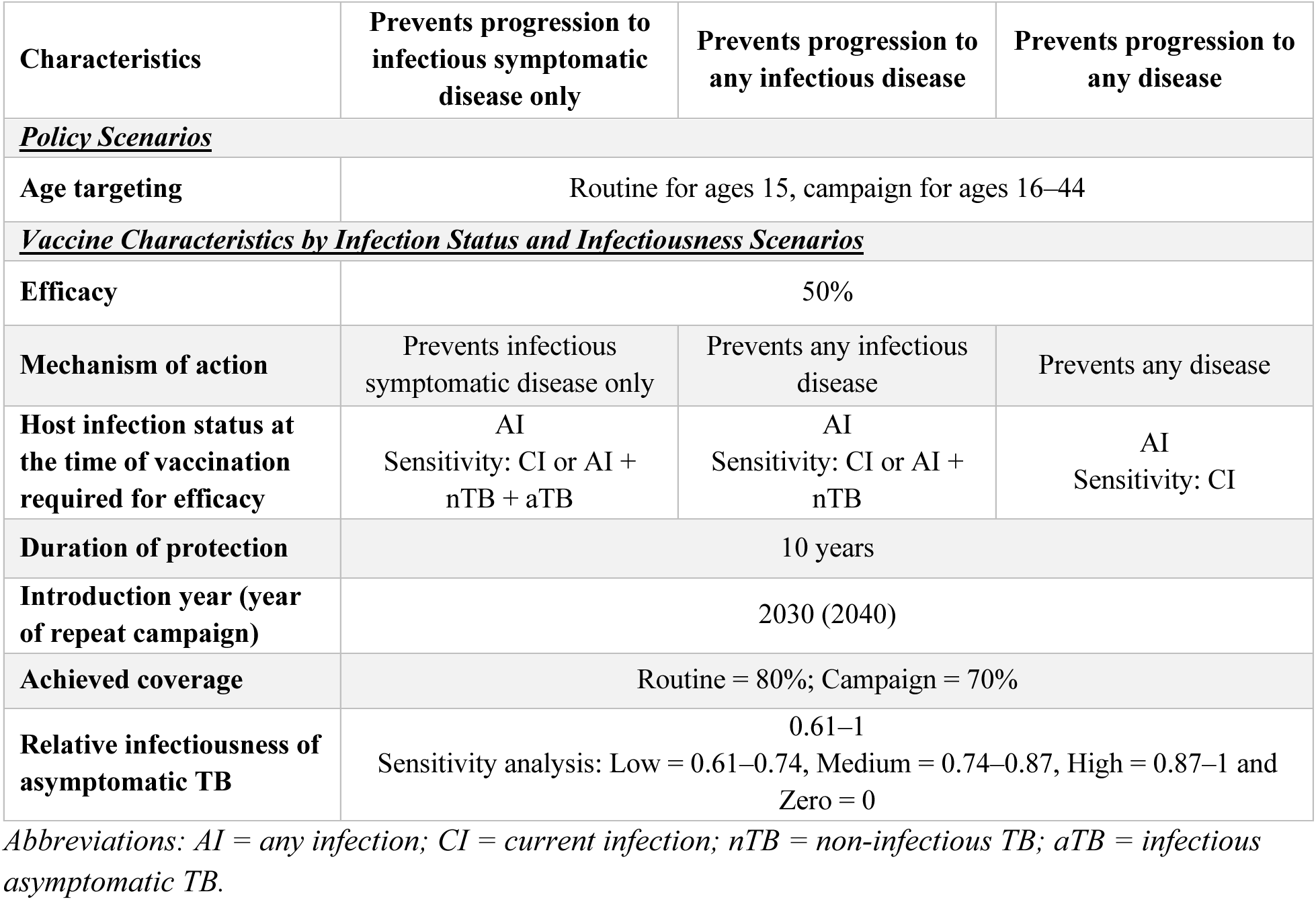
New TB vaccine policy scenario characteristics.

| Characteristics | Prevents progression to infectious symptomatic disease only | Prevents progression to any infectious disease | Prevents progression to any disease |
| --- | --- | --- | --- |
| ***<u>Policy Scenarios</u>*** |  |  |  |
| Age targeting | Routine for ages 15, campaign for ages 16–44 |  |  |
| ***<u>Vaccine Characteristics by Infection Status and Infectiousness Scenarios</u>*** |  |  |  |
| Efficacy | 50% |  |  |
| Mechanism of action | Prevents infectious symptomatic disease only | Prevents any infectious disease | Prevents any disease |
| Host infection status at the time of vaccination required for efficacy | AI<br>Sensitivity: CI or AI + nTB + aTB | AI<br>Sensitivity: CI or AI + nTB | AI<br>Sensitivity: CI |
| Duration of protection | 10 years |  |  |
| Introduction year (year of repeat campaign) | 2030 (2040) |  |  |
| Achieved coverage | Routine = 80%; Campaign = 70% |  |  |
| Relative infectiousness of asymptomatic TB | 0.61–1<br>Sensitivity analysis: Low = 0.61–0.74, Medium = 0.74–0.87, High = 0.87–1 and Zero = 0 |  |  |

*Abbreviations: AI = any infection; CI = current infection; nTB = non-infectious TB; aTB = infectious asymptomatic TB.*

We simulated three ways the vaccine could affect progression to disease (i.e., the mechanisms of action) (12):

1. Preventing progression to *infectious symptomatic disease only* (red dashed line in Figure 1b).
2. Preventing progression to *any infectious disease* (blue dash-dotted lines in Figure 1b)
3. Preventing progression to *any disease* (grey dashed lines in Figure 1b)

### Sensitivity analyses

We analysed the sensitivity of our results to a) the host infection status at time of vaccination required for efficacy, b) the relative infectiousness of aTB, and c) whether the vaccine also protects if given to individuals with disease at the time of vaccination. For (a) we instead assumed that only individuals with infection at time of vaccination could be protected (i.e., modelling a ‘current infection’ vaccine), aligning with the evidence from the M72/AS01_E_ Phase 2b trial (12). For (b), four scenarios of the relative infectiousness were modelled: three scenarios created by dividing the full range of relative infectiousness into low (0.61–0.74), medium (0.74–0.87), and high (0.87–1) categories, and a scenario with zero infectiousness, and the model was recalibrated. For (c), we instead assumed vaccines were also effective in those with pre-symptomatic disease at time of vaccination (Figure 1c). See supporting methods for full details.

### Outcomes

We calculated the number and proportion of sTB episodes, aTB episodes, nTB episodes, and TB deaths averted between 2030–2050. We show the short-term impact between 2030–2032 which primarily captures the vaccine direct effects and is a rough approximation to the duration of a typical TB vaccine trial, and the longer-term impact which captures the direct and indirect effects over 20 years.

## Results

### No-new-vaccine scenario results

The *no-new-vaccine* scenario fit all 14 calibration targets with 500 parameter sets (Supplementary Table S 5). Over 2030–2050, the *no-new-vaccine* scenario predicted 47.0 million (40.4, 56.2) incident sTB episodes, 175.8 million (136.4, 216.3) incident aTB episodes, 233.9 million (166.5, 324.6) incident nTB episodes, and 8.8 million (7.5, 10.7) TB deaths (Figure S 5).

### New TB vaccine policy scenario results

The short-(2030–2032) and longer-term (2030–2050) impact of vaccines with 50% efficacy to prevent disease delivered routinely to 15-year-olds and as two campaigns to those aged 16–44 and effective with any infection status at the time of delivery (the *Basecase* scenarios) are shown in Figure 2 (percent averted) and Table 2 (percent and numbers averted).

**Figure 2.**
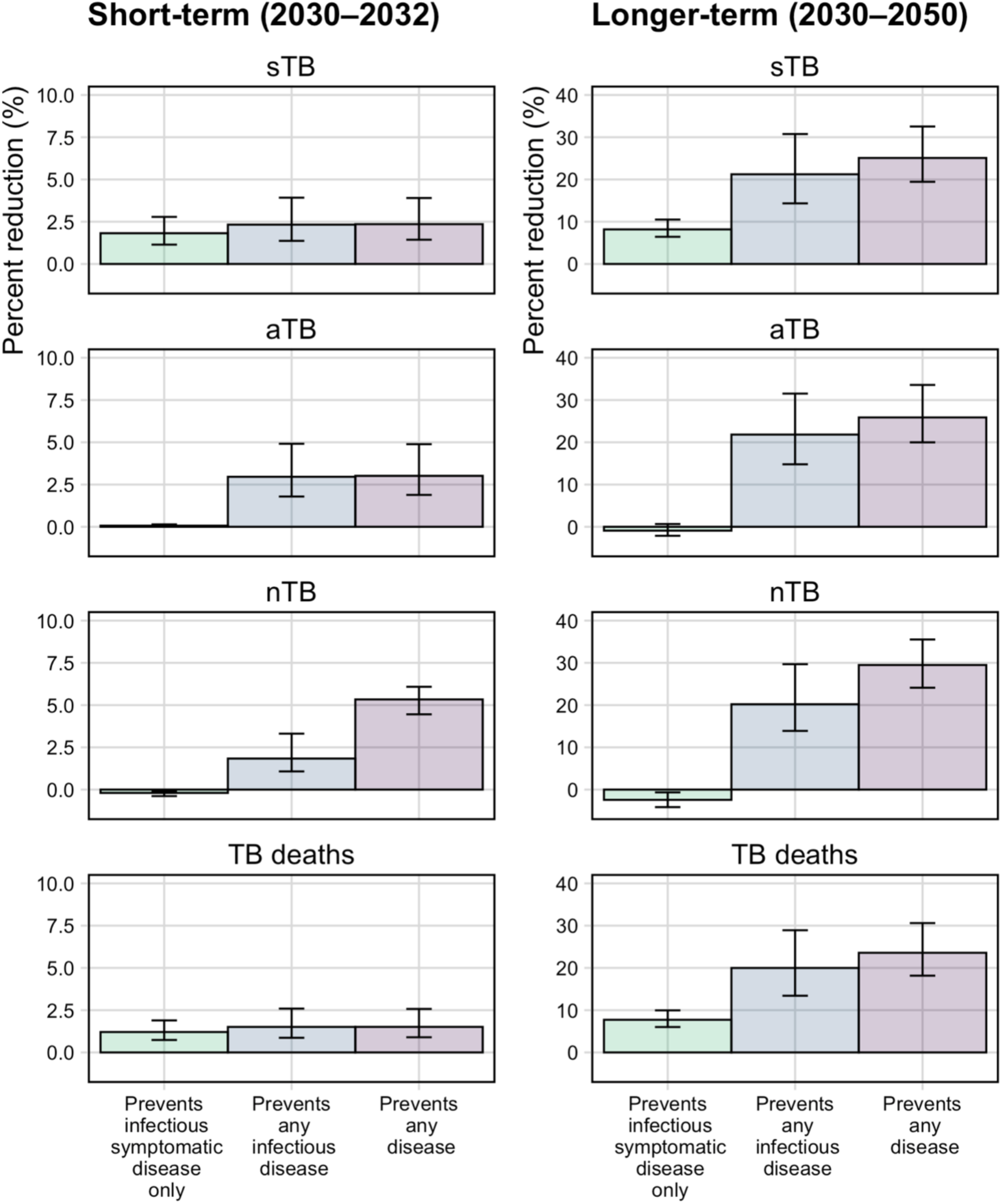
Percentage reduction in infectious symptomatic TB episodes (sTB, top row), infectious asymptomatic TB episodes (aTB, second row), non-infectious TB episodes (nTB, third row), and TB deaths (bottom row) over short-(2030–2032, left column) and longer-term (2030–2050, right column), by vaccine disease prevention mechanism of action (preventing infectious symptomatic disease only, any infectious disease, or any disease). *Note y-axis scales differ between short and longer term*.

**Table 2.** Number and cumulative percentage of infectious symptomatic TB (sTB), infectious asymptomatic TB (aTB), infectious symptomatic and infectious asymptomatic TB (aTB + sTB), non-infectious TB (nTB) episodes, and TB deaths averted over short- (2030–2023) and longer-term (2030–2050), by vaccine disease prevention mechanism of action (infectious symptomatic disease only, any infectious disease or any disease).

|  |  | Short-term (2030–2032) |  |  | Longer-term (2030–2050) |  |  |
| --- | --- | --- | --- | --- | --- | --- | --- |
| Vaccine prevents progression to>> |  | Infectious symptomatic disease only | Any infectious disease | Any disease | Infectious symptomatic disease only | Any infectious disease | Any disease |
| Numbers averted (millions) | sTB | 0.13<br>(0.08, 0.21) | 0.17<br>(0.09, 0.29) | 0.17<br>(0.10, 0.28) | 3.83<br>(2.67, 5.58) | 9.87<br>(6.22, 15.54) | 11.81<br>(8.20, 16.61) |
|  | aTB | 0.02<br>(0.01, 0.04) | 0.79<br>(0.45, 1.42) | 0.80<br>(0.47, 1.40) | -1.55<br>(-3.77, 1.15) | 37.79<br>(22.47, 61.69) | 45.26<br>(29.83, 66.27) |
|  | aTB + sTB | 0.15<br>(0.09, 0.24) | 0.95<br>(0.54, 1.71) | 0.97<br>(0.57, 1.70) | 2.26<br>(-0.26, 6.06) | 47.64<br>(28.81, 76.68) | 57.23<br>(38.36, 82.71) |
|  | nTB | -0.07<br>(-0.12, -0.04) | 0.64<br>(0.36, 1.06) | 1.88<br>(1.27, 2.61) | -5.70<br>(-9.58, -1.56) | 47.57<br>(28.50, 69.46) | 69.42<br>(44.66, 96.51) |
|  | TB deaths | 0.02<br>(0.01, 0.03) | 0.02<br>(0.01, 0.04) | 0.02<br>(0.01, 0.04) | 0.69<br>(0.48, 0.98) | 1.77<br>(1.10, 2.77) | 2.11<br>(1.47, 2.99) |
| Percent reduction (%) | sTB | 1.82%<br>(1.14, 2.79) | 2.33%<br>(1.37, 3.92) | 2.36%<br>(1.44, 3.90) | 8.19%<br>(6.43, 10.51) | 21.23%<br>(14.35, 30.76) | 25.09%<br>(19.45, 32.54) |
|  | aTB | 0.07%<br>(0.03, 0.14) | 2.96%<br>(1.79, 4.91) | 3.02%<br>(1.89, 4.89) | - 0.91%<br>(-2.13, 0.65) | 21.83%<br>(14.78, 31.53) | 25.88%<br>(19.99, 33.58) |
|  | aTB + sTB | 0.44%<br>(0.27, 0.69) | 2.83%<br>(1.71, 4.72) | 2.88%<br>(1.80, 4.69) | 1.01%<br>(-0.13, 2.62) | 21.70%<br>(14.68, 31.38) | 25.70%<br>(19.87, 33.36) |
|  | nTB | -0.20%<br>(-0.39, -0.10) | 1.84%<br>(1.08, 3.31) | 5.33%<br>(4.45, 6.08) | -2.43<br>(-4.15, -0.65) | 20.20%<br>(13.89, 29.67) | 29.48%<br>(24.11, 35.52) |
|  | TB deaths | 1.21%<br>(0.74, 1.89) | 1.51%<br>(0.87, 2.59) | 1.53%<br>(0.90, 2.57) | 7.73%<br>(6.01, 9.94) | 19.99%<br>(13.41, 28.91) | 23.58%<br>(18.16, 30.61) |

### Impact on infectious symptomatic TB episodes (sTB)

In the short-term, there was little difference in the proportion of cumulative sTB episodes between vaccines preventing progression to only infectious symptomatic disease, any infectious disease or any disease (1.8% [1.1, 2.8], 2.3% [1.3, 3.9] and 2.4% [1.4, 3.9], respectively) compared to the *no-new-vaccine* scenario (Figure 2, Table 2).

However, over the longer-term, compared to a vaccine that prevented only infectious symptomatic disease, vaccines preventing progression to any infectious disease or any disease averted a much larger proportion of sTB episodes (8.2% [6.4, 10.5], 21.0% [14.4, 30.8], and 25.1% [19.5, 32.5], respectively) (Figure 2, Table 2), due to preventing continued transmission from aTB.

### Impact on infectious asymptomatic TB episodes (aTB)

In the short-term, all three vaccine types averted aTB episodes compared to the *no-new-vaccine* scenario, with reductions of 0.1% (0.0, 0.14), 3.0% (1.8, 4.9), and 3.0% (1.9, 4.9) for vaccines preventing infectious symptomatic disease only, any infectious disease, and any disease respectively (Figure 2, Table 2).

Over the longer-term, only vaccines preventing any infectious disease or any disease averted episodes of aTB compared to the *no-new-vaccine* scenario (21.9% [14.8, 31.5] and 25.9% [20.0, 33.6], respectively), whilst there was a small (0.8% [0.6, 2.1]) *increase* in episodes for a vaccine only preventing infectious symptomatic disease compared to the *no-new-vaccine* scenario (Figure 2, Table 2). The increase in aTB episodes for a vaccine only preventing infectious symptomatic TB was observed due to the accumulation of individuals in aTB contributing to transmission.

### Impact on non-infectious TB episodes (nTB)

In the short-term, there was a small (0.2% [0.1, 0.4]) *increase* in nTB episodes for a vaccine that prevented infectious symptomatic disease only, and vaccines that prevented any infectious disease or any disease averted 1.8% (1.1, 3.3) and 5.3% (4.5, 6.1) of nTB episodes, respectively, compared to the *no-new-vaccine* scenario (Figure 2, Table 2).

Over the longer-term, vaccines that prevented any infectious disease or any disease averted 20.2% (13.9, 29.7) and 29.5% (24.1, 35.5) and of nTB episodes, respectively, compared to the *no-new-vaccine* scenario, whilst there was a small (2.4% [0.6, 4.1]) *increase* in nTB episodes for a vaccine that only prevented infectious symptomatic disease (Figure 2, Table 2). This is because the vaccine only preventing infectious symptomatic disease allows individuals to accumulate and remain in the aTB stage for longer, contributing to greater transmission overall and increasing the number of nTB episodes observed.

### Impact on TB deaths

In the short-term, there was little difference in the proportion of cumulative TB deaths averted by vaccines that prevented only infectious symptomatic disease, any infectious disease, or any disease (1.2% [0.7, 1.9], 1.5% [0.9, 2.6], and 1.5% [0.9, 2.6], respectively).

However, over the longer-term, compared to a vaccine that prevented only infectious symptomatic TB, vaccines preventing any infectious disease or any disease averted a much larger proportion of cumulative TB deaths (7.7% [6.0, 9.9], 20.0% [13.4, 28.9], and 23.6% [18.2, 30.6], respectively) (Figure 2, Table 2), again due to the reduction of transmission by preventing aTB episodes.

### Trends over time

Figure 3 shows trends in the number of sTB episodes, aTB episodes, nTB episodes, and TB deaths for the *no-new-vaccine* and each *Basecase* scenario over time from 2030–2050. The figure shows that a vaccine preventing only infectious symptomatic disease may *increase* the number of nTB and aTB episodes compared to the *no-new-vaccine* scenario (green trend line vs yellow trend line). A vaccine preventing only infectious symptomatic disease may initially have marked impact on sTB episodes and TB deaths, but this impact may reduce over time as the individuals with aTB accumulate and continue to transmit.

**Figure 3.**
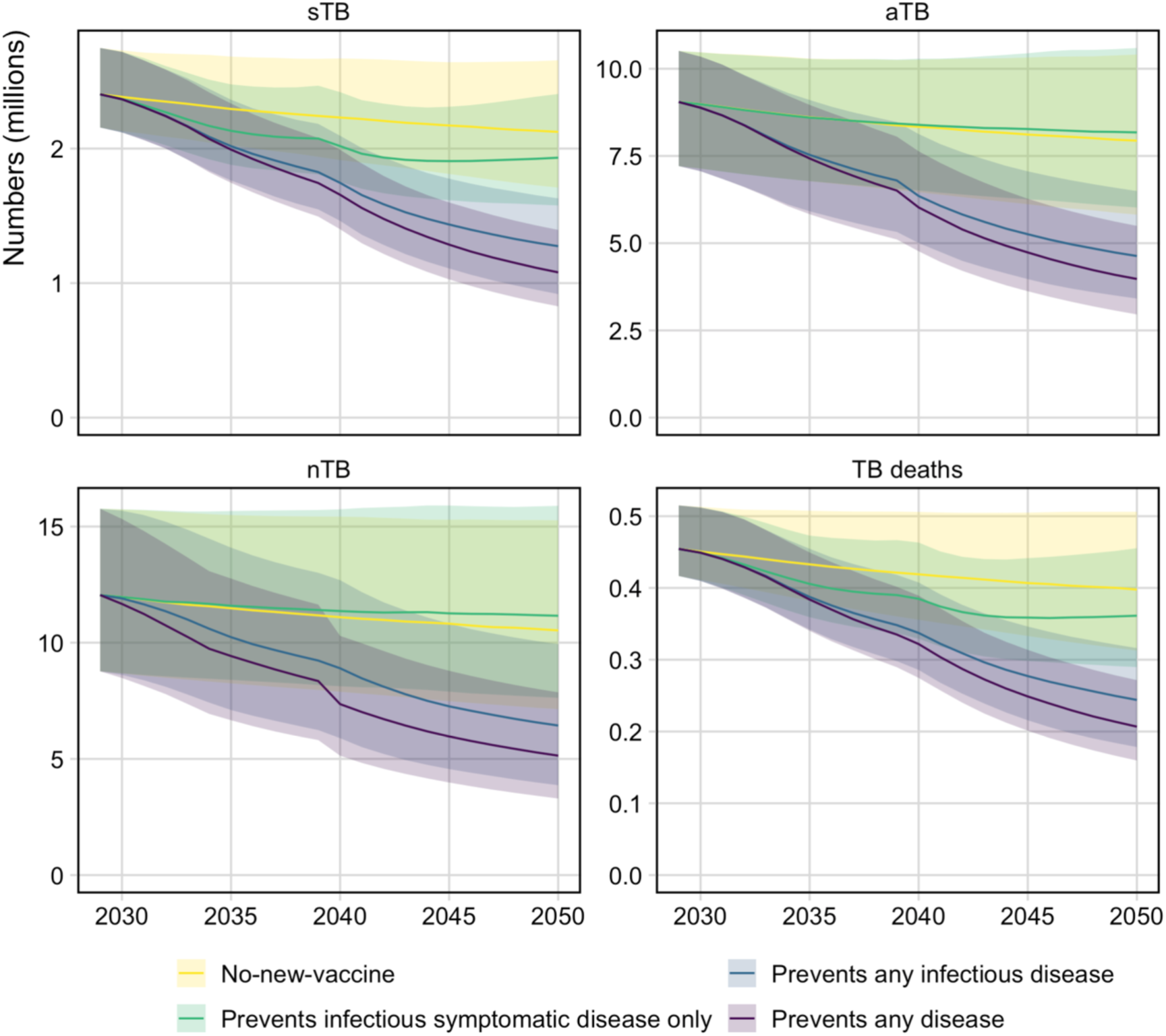
Trends in the number of sTB episodes, aTB episodes, nTB episodes and TB deaths between 2030–2050 for each vaccine scenario compared to no-new-vaccine scenario. *Note y-axis scales differ*.

### Sensitivity analyses

*Efficacy by host infection status at the time of vaccination required for vaccine efficacy* If the vaccine was only effective in those with current infection at the time of vaccination, as expected, we saw lower impact from all vaccines and at all time points compared to the *Basecase* scenarios. However, the patterns and trends with respect to the mechanism of action of the vaccine were similar to those observed for the *Basecase* scenarios (Figure S 8, Table S 8).

### Relative infectiousness of aTB

When the relative infectiousness of aTB compared to sTB was varied within the range of current literature estimates (0.61–1), short- and longer-term results were similar to the *Basecase* scenario impact estimates (Figure S 10, Table S 9, Figure S 11, Table S 10). Under the assumption that aTB was not infectious at all, results remained similar to the *Basecase* scenarios in the short-term but differed in the longer-term (Figure S 10, Table S 9, Figure S 11, Table S 10) because there was no indirect transmission effect of preventing aTB episodes.

### Efficacy if given to individuals with nTB or aTB

Supporting Figure S 12 and Table S 11 show the short-term vs. longer-term vaccine impact if an ‘any infection’ or ‘current infection’ vaccine was also effective in early disease stages. Results suggest a vaccine that prevented any infectious disease had greater impact for all outcomes, both short- and longer-term, when effective in individuals with nTB at vaccination, compared to when the vaccine was ineffective if given to those individuals. In contrast, a vaccine that prevented only infectious symptomatic disease had a more pronounced impact when effective in individuals with nTB or aTB, leading to more nTB episodes and greater reductions in sTB episodes and deaths, compared to when ineffective in individuals with nTB or aTB. See supporting results section 10 for full details.

## Discussion

This is the first modelling study to investigate the sensitivity of impact estimates of new TB vaccines to uncertainties in vaccine efficacy by TB disease stage and aTB infectiousness.

Results suggest that over a three-year period—similar to the duration of a typical vaccine trial— vaccines that prevented only infectious symptomatic disease, any infectious disease, or any form of TB averted a similar proportion of cumulative sTB episodes. However, over a 20-year period, which better reflects the full public health impact of vaccination, the differences became more pronounced. Compared to a vaccine that only prevented infectious symptomatic TB, vaccines preventing any infectious disease or any disease averted a significantly higher proportion of sTB episodes. This increased impact was due to preventing the accumulation of and ongoing transmission from aTB. Although the effect was small, a vaccine targeting only infectious symptomatic TB also led to an *increase* in aTB and nTB episodes. These findings highlight the vaccine’s mechanism of action: by preventing only infectious symptomatic disease, this resulted in a growing pool of individuals with aTB, sustaining transmission over time.

We explored how the vaccine impact could be affected if we were wrong in some of our key assumptions. In general, our results were robust to our assumptions, but they were sensitive to zero infectiousness of aTB, and if vaccine efficacy was assumed in early pre-symptomatic disease stages. If aTB was assumed to have zero infectiousness, then the differences in impact between vaccine types on preventing sTB episodes was reduced. Over the longer-term, all vaccine types averted a similar proportion of sTB episodes, as accumulation of individuals in the aTB stage under the scenario where the vaccine only prevented progression to symptomatic disease would not result in continued transmission from aTB. However, in support of the assumption used in the *Basecase* analyses that the infectiousness of aTB relative to sTB was 0.61–1, although limited in number, studies have suggested that aTB *is* likely to be infectious and contribute to transmission (2,30). We also evaluated scenarios where we assumed, hypothetically, that the vaccines would be effective in pre-symptomatic disease stages at the time of vaccination. If this was the case, we saw greater impact from the vaccines due to protecting a larger proportion of the population, specifically those who were at a high risk of progressing to further disease stages. However, experts believe that it is unlikely that a vaccine would be effective if delivered to someone with disease at the time of vaccination, as the immune response would overwhelm any vaccine effect. Although, whether the same applies for earlier disease stages (such as nTB) and undulation between disease stages is not yet known.

Our work has limitations. We modelled the impact of vaccine scenarios designed to prevent only infectious symptomatic disease only, any infectious disease, or any disease in India, and these findings may differ in other settings. We assumed that coverage of interventions post-2022 would remain constant. If future non-vaccine interventions were scaled up, we will have overestimated absolute vaccine impacts, but relative impacts may be similar. Estimates of vaccine efficacy and duration of protection were based on trial data and expert input, which may not reflect realised efficacy. We assumed the vaccine would be equally effective in preventing all disease stages. If the vaccine is more or less effective by disease stage, our model may under or overestimate impact. We took a maximalist posture on scale-up of vaccine coverage to illustrate the potential impact, but real-world supply constraints and introduction decisions would likely mitigate the realised impact. Our results depend on natural history structures and initial parameter ranges from established literature reviews, which may not accurately represent the dynamics of TB disease. We did not account for the effect of comorbidities such as HIV, undernutrition, or diabetes on disease transmission and progression. Therefore, the impact of vaccines under these conditions could differ. Finally, certain modelling assumptions, such as the exclusion of mortality from nTB, represent further simplifications that may impact our projections. As nTB may represent individuals with extra-pulmonary TB, by assuming no extra nTB mortality, we may be underestimating the benefits of early TB stage vaccine efficacy (31). These limitations could be investigated in future work.

There are currently around 15 vaccine candidates in trials and the average follow-up time for prevention of disease trial participants is around three years (32). The primary endpoint of these trials is typically sTB, with impact on aTB either not being measured, or being measured as a secondary and potentially underpowered endpoint. Therefore, at the time of licensure, we may only have powered estimates on the impact of new vaccines on sTB. By having trial data only on the short-term impact on sTB episodes, we will be unable to determine which type of TB disease the vaccine prevents and will not have the data needed to estimate the full public health value of the vaccine. One solution could be to carry out Phase 4 studies after licensure, which may help us know if the wider population level impact is lower than expected. However, this would take many years after licensure for results and would be confounded by other factors such as vaccine coverage and the roll-out of other non-vaccine strategies. Alternatively, a quicker and more robust solution could be to separately measure impact on both aTB and sTB in Phase 2b and 3 trials. If the effect on aTB was measured, for example, by collecting sputum on all participants during or at the end of trials (5), impact by TB disease stage could be known alongside vaccine licensure and would provide critical evidence on the vaccine mechanism of action for global and country vaccine decision makers considering vaccine roll out.

In addition, empirical data should be collected to better characterise aTB. There are currently limited direct evidence on morbidity, infectiousness, and progression to sTB (33,34). Future data collection could not only help to better estimate the potential full value of new TB vaccines, but also to better understand the clinical and public health consequences of aTB.

If the assumptions on the characteristics of aTB hold, information from this study may be important. WHO may revise their Preferred Product Characteristics for New TB Vaccines (35) to specify that vaccines should also prevent earlier stages of disease. Vaccine developers may prioritise development of vaccines that also prevent earlier stages of TB disease. TB vaccine trialists may want to test efficacy against aTB to provide timely information about the mechanism of action (5). It is important to note that a vaccine that only prevents infectious symptomatic disease is still likely to have a large public health benefit on TB morbidity and mortality. However, other programmatic efforts may be needed to avoid the accumulation of and transmission from individuals with infectious asymptomatic disease.

The Full Value of Vaccine Assessment framework incorporates a wide range of outcomes, both health and non-health (societal/economic), direct and indirect, and to the individual or the population (36). This study focussed on the traditional direct health impacts of introducing new TB vaccines, but future work could investigate the wider economic impacts of vaccines with varying efficacy by disease stage and estimate whether vaccines that prevent only infectious symptomatic disease could be cost-effective given reductions in TB deaths, treatment costs and loss of productivity.

With our evolving understanding of TB natural history and WHO recognising the potential importance of aTB, we should be open to course-correct, from trial design to estimates of vaccine impact. The population impact of new TB vaccines may depend on efficacy against infectious asymptomatic TB. TB vaccine trials should measure impact on aTB to enable better estimates of the potential full value of new TB vaccines. Further data collection is required to better understand the transmissibility, morbidity, and dynamics of asymptomatic disease.

## Data Availability

No individual level participant data was used for this modelling study. Analytic code will be made available immediately following publication indefinitely for anyone who wishes to access the data for any purpose.

