## Supplementary Material for "Population impact of new TB vaccines may depend on efficacy against infectious asymptomatic TB: a modelling study"

### Table of Contents

**SUPPORTING METHODS** **2**

1. MODEL STRUCTURE AND EQUATIONS 2
   - *1.1 Natural history model structure 2*
   - *1.2 Natural history scenarios 3*
   - *1.3 Model equations 4*
2. MODEL PARAMETERS AND DATA SOURCES 5
   - *2.1 Natural history parameter values and data sources 5*
   - *2.2 Age varying parameters 8*
   - *2.3 Treatment initiation and outcomes 9*
3. MODEL SIMULATION AND CALIBRATION 11
   - *3.1 Model simulation 11*
   - *3.2 Model calibration 11*
4. POLICY SCENARIOS 13
   - *4.1 No-new-vaccine scenario 13*
   - *4.2 Vaccine scenarios 13*
   - *Vaccine integration in the TB natural history model 16*

**SUPPORTING RESULTS** **18**

5. NO-NEW-VACCINE SCENARIO RESULTS 18
   - *5.1 No-new-vaccine calibration (with baseline, low, medium and high relative infectiousness) 18*
   - *5.2 No-new-vaccine scenario calibration (with zero relative infectiousness) 19*
6. VACCINATED PROPORTIONS OVER TIME 21
7. SENSITIVITY ANALYSIS RESULTS: VACCINES EFFECTIVE WITH CURRENT INFECTION STATUS 22
8. TRENDS OVER TIME FOR VACCINES EFFECTIVE WITH CURRENT INFECTION STATUS 24
9. SENSITIVITY ANALYSIS RESULTS: VACCINES EFFECTIVE WITH ANY INFECTION STATUS WITH VARYING INFECTIOUSNESS 25
   - *9.1 Short-term impact (2030–2032) 25*
   - *9.2 Longer-term impact (2030–2050) 29*
10. SENSITIVITY ANALYSIS RESULTS: VACCINES EFFECTIVE WITH ANY OR CURRENT INFECTION STATUS INCLUDING EFFICACY IN PRE-DISEASE STAGES 31

**SUPPORTING DISCUSSION** **34**

### SUPPORTING METHODS

### 1. Model structure and equations

We created an age-stratified compartmental differential equation model of tuberculosis in India, including dimensions for age, tuberculosis natural history, vaccination, and treatment. The age structure is identical to that included in Clark et al., *BMC Medicine*, 2023 (1). The natural history structure is an adaptation from Clark et al. informed by work on the natural history of TB from Richards et al., *Lancet Glob Health*, 2023 (2) and Horton et al., *PNAS*, 2023 (3). Symptomatic disease compartment naming has been done adhering to current TB definitions according to the latest WHO guidance, and definitions of asymptomatic TB following a WHO consultation 2024 (4).

#### 1.1 Natural history model structure

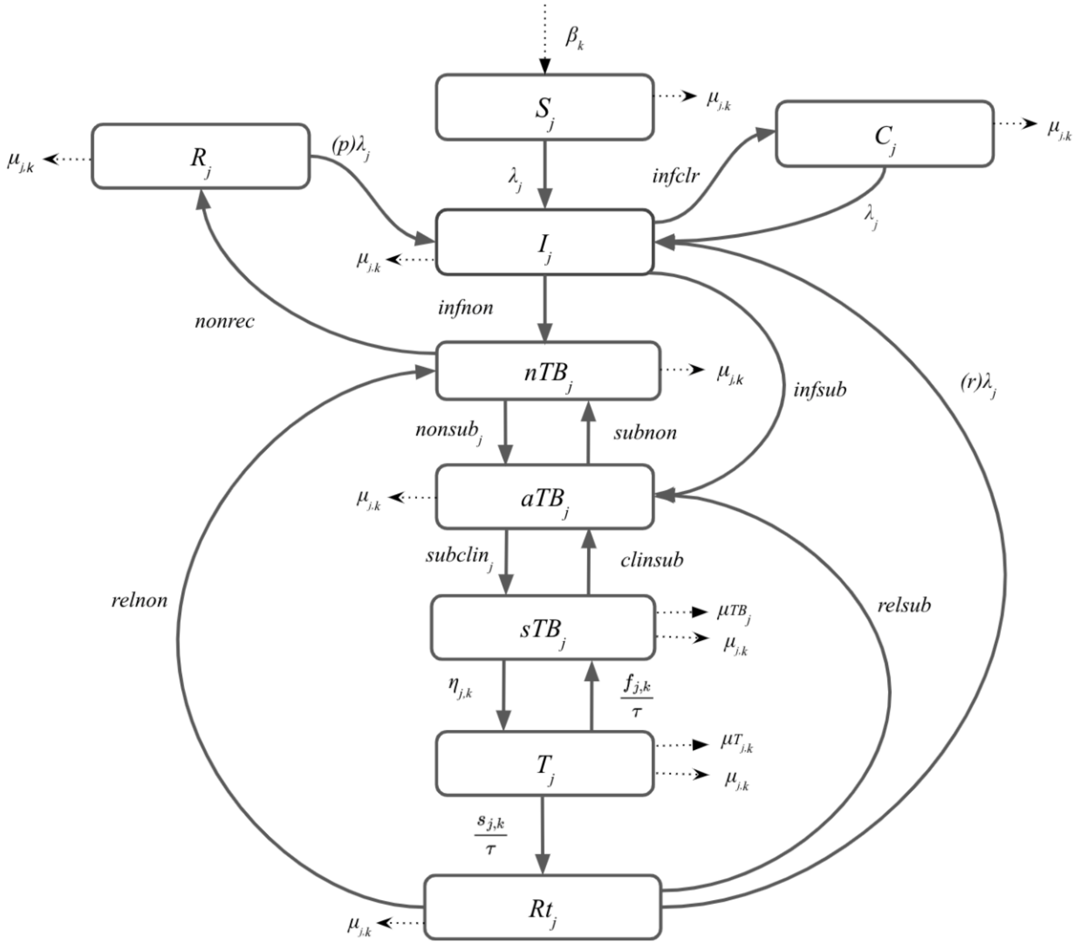


**Figure S 1 Tuberculosis natural history model structure**

*Abbreviations: S = Susceptible; I = Infection; C = Cleared; nTB = Non-infectious TB; R = Recovered; aTB = Infectious asymptomatic TB; sTB = Infectious symptomatic TB; T = On-Treatment; Rt = Recovered after treatment. Subscript j represents parameters that vary by age, and subscript k represents parameters that vary over time.*

A natural history structure with nine compartments in Figure S 1 was created by adapting features of previous models.

- Susceptible individuals (S) are infected at rate $\lambda$ and progress to the infected compartment. Infected individuals $(I)$ may clear infection $(C)$ at rate $infclr$ or progress to non-infectious disease $(nTB)$ or infectious asymptomatic disease $(aTB)$ at rates $infnon$ and $infsub$, respectively.
- Individuals with non-infectious disease $(nTB)$ recover $(R)$ at rate $nonrec$ or progress to infectious asymptomatic disease $(aTB)$ at rate $nonsub$. Individuals with infectious asymptomatic disease $(aTB)$ regress to non-infectious disease $(nTB)$ at rate $subnon$ or progress to infectious symptomatic disease $(sTB)$ at rate $subclin$. Individuals with infectious symptomatic disease $(sTB)$ regress to infectious asymptomatic disease $(aTB)$ at rate $clinsub$ or die from TB-associated mortality at rate $\mu TB$.
- Individuals with infectious symptomatic disease $(sTB)$ progress to on-treatment $(T)$ at rate $\eta$. Individuals on-treatment $(T)$ have three possible treatment outcomes, whereby individuals can progress to recovered after treatment $(Rt)$ at on-treatment completion rate $\frac{s_j}{\tau}$, individuals can regress to infectious symptomatic disease $(sTB)$ at on-treatment non-completion rate $\frac{f_j}{\tau}$, or die while on-treatment.
- Individuals recovered after treatment $(Rt)$ may relapse to non-infectious disease $(nTB)$ or to infectious asymptomatic disease $(aTB)$ at rates $relnon$ and $relsub$, respectively.
- Individuals who have recovered $(R)$ from non-infectious disease $(nTB)$ are protected from reinfection with an effective reinfection rate of $p\lambda$ and those who have recovered after treatment $(Rt)$ have an increased risk of reinfection $(r\lambda)$.

#### 1.2 Natural history scenarios

We explored the impact of varying the infectiousness of aTB relative to sTB on the impact of TB vaccines by dividing the baseline range of aTB infectiousness relative to sTB (0.61–1) into three equal segments: low (0.61–0.74), medium (0.74–0.87), and high (0.87–1) infectiousness. We then calibrated the model to reflect all four scenarios—baseline, low, medium, and high—and compared the effect of the infectiousness scenarios on the resulting impact of TB vaccines. We also investigated a scenario where relative aTB infectiousness was zero.

### 1.3 Model equations

#### Natural history model equations

$$\frac{dS_j}{dt} = \beta_k - (\lambda_j + \mu_{j,k})S_j \ where \ Age_j = 0$$

$$\frac{dS_j}{dt} = -(\lambda_j + \mu_{j,k})S_j \ where \ Age_j \neq 0$$

$$\frac{dC_j}{dt} = infclr(I_j) - (\lambda_j + \mu_{j,k})C_j$$

$$\frac{dR_j}{dt} = nonrec(nTB_j) - (p\lambda_j + \mu_{j,k})R_j$$

$$\frac{dI_j}{dt} = \lambda_j(S_j + C_j + pR_j + rRt_j) - (infclr + infnon + infsub + \mu_{j,k})I_j$$

$$\frac{dnTB_j}{dt} = infnon(I_j) + relnon(Rt_j) + subnon(aTB_j) - (nonrec + nonsub_j + \mu_{j,k})nTB_j$$

$$\frac{daTB_j}{dt} = infsub(I_j) + nonsub_j(nTB_j) + relsub(Rt_j) + clinsub(sTB_j) - (subnon + subclin_j + \mu_{j,k})aTB_j$$

$$\frac{dsTB_j}{dt} = subclin_j(aTB_j) + \frac{f_{j,k}}{\tau}(T_j) - \left(clinsub + \eta_{j,k} + \mu_{TB_j} + \mu_{j,k}\right)sTB_j$$

$$\frac{dT_j}{dt} = \eta_{j,k}(sTB_j) - \left(\frac{s_{j,k} + f_{j,k}}{\tau} + \mu_{T\,j,k} + \mu_{j,k}\right)T_j$$

$$\frac{dRt_j}{dt} = \frac{s_{j,k}}{\tau}(T_j) - (relnon + relsub + r\lambda_j + \mu_{j,k})Rt_j$$

**Force of infection equation**

Susceptible individuals (S) are infected at rate $\lambda$. $\lambda$ is defined as follows:

$$\lambda_j = pT \cdot \sum_{y=1}^{n_{ygroups}} C[m,y] \cdot \left( \frac{(1-ep) \cdot \left( T_{sTB_y} + tT_{aTB_y} \right) \cdot k_{inf}}{N_y} \right)$$

| | |
|---|---|
| $j$ | indicates age of individual in years |
| $pT$ | indicates probability of transmission per infectious contact |
| $n_{groups}$ | indicates the number of contact age groups |
| $C[m,y]$ | indicates the number of age-specific contacts |
| $m$ | indicates age group of individual |
| $y$ | indicates the age group of contact |
| $T_{sTB_y}$ | indicates the total number of individuals with symptomatic TB in age group $y$ |
| $T_{aTB_y}$ | indicates the total number of individuals with asymptomatic TB in age group $y$ |
| $k_{inf}$ | indicates the relative infectiousness of children (<15 years) relative to adults (≥15 years) |
| $ep$ | indicates the average proportion of TB cases that are extrapulmonary |
| $t$ | indicates the infectiousness of asymptomatic TB relative to symptomatic TB |
| $N_y$ | indicates the total population in age group $y$ |

### 2. Model Parameters and Data Sources

#### 2.1 Natural history parameter values and data sources

Table S 1 shows the India model parameters and sources used in the natural history model structure, along with their definitions, sources, and information on whether the parameter was fixed or varied (as well as whether they were varied by age or time) during calibration.

Further details about how the age varying parameters were implemented are provided in section 2.2, and further details related to TB treatment are provided in section 2.3. The parameter ranges provided for the TB natural history parameters are priors used during calibration in a Bayesian analysis. We assumed that all values within the prior range were equally likely. The prior ranges were pre-specified based on literature review and were reviewed as new data became available.

**Table S 1 India national model parameter values and sources**

| Description | Units | Symbol | Initial Range | Fixed or Varying During Calibration | Age Varying | Time Varying | Source |
|---|---|---|---|---|---|---|---|
| ***Births and deaths (excluding on-treatment mortality)*** | | | | | | | |
| Birth rate | Per year | $\beta_k$ | UN World Population Prospects population estimates and projections | Fixed | No | Yes | (5) |
| Background mortality rate | Per year | $\mu_{j,k}$ | Calculated in the model from UN population estimates and projections | Fixed | Yes, age specific mortality rates from demographic dataset | Yes | (5) |
| Mortality rate for $sTB$ | Per person per year | $\mu TB_j$ | (0.28–0.38) | Varying | Yes, value for children is greater than value for adults | No | (3) |
| ***Natural History*** | | | | | | | |
| Force of infection | Per year | $\lambda_j$ | Fitted | Fixed Equation | Yes, age specific contact rates(6) | No | *Calculated* |
| Probability of transmission per infectious contact | - | $pT$ | (0–0.0068) | Varying | No | No | *Assumed* |
| Fraction of total TB that is extrapulmonary | - | $ep$ | 0.1972 | Fixed | No | No | (7,8) |
| Infectiousness of $aTB$ relative to $sTB$ | - | $t$ | (0.61–1) | Varying | No | No | (9) |
| Rate from $I$ to $aTB$ | Per person per year | $infsub$ | (0.01–0.1) | Varying | No | No | (3) |
| Rate from $I$ to $nTB$ | Per person per year | $infnon$ | (0.04–0.23) | Varying | No | No | (3) |
| Rate from $I$ to $C$ | Per person per year | $infclr$ | (0.93–3.30) | Varying | No | No | (3) |
| Rate from $nTB$ to $R$ | Per person per year | $nonrec$ | (0.14–0.23) | Varying | No | No | (3) |

| | | | | | | | |
|---|---|---|---|---|---|---|---|
| Rate from $nTB$ to $aTB$ | Per person per year | $nonsub_j$ | (0.21–0.28) | Varying | Yes; value for children is **less** than value for adults. | No | (3) |
| Rate from $aTB$ to $nTB$ | Per person per year | $subnon$ | (1.24–2.03) | Varying | No | No | (3) |
| Rate from $aTB$ to $sTB$ | Per person per year | $subclin_j$ | (0.56–0.94) | Varying | Yes; value for children is **less** than value for adults. | No | (3) |
| Rate from $sTB$ to $aTB$ | Per person per year | $clinsub$ | (0.46–0.72) | Varying | No | No | (3) |
| Rate of relapse from $Rt$ to $nTB$ | Per person per year | $relnon$ | (0–0.01) | Varying | No | No | *Assumed* |
| Rate of relapse from $Rt$ to $aTB$ | Per person per year | $relsub$ | (0–0.01) | Varying | No | No | *Assumed* |
| ***Protection Parameters*** | | | | | | | |
| Relative risk of re-infection in $R$ | - | $p$ | (0.14–0.30) | Varying | No | No | (10) |
| Relative risk of re-infection in $Rt$ | - | $r$ | (2.14–4.27) | Varying | No | No | (11) |
| ***Treatment parameters*** | | | | | | | |
| Treatment initiation from $sTB$ | Per person per year | $\eta_{j,k}$ | 0–1 | Sigmoidal curve describing rate of treatment initiation | Yes; value for children is **less** than value for adults. | Yes | *Assumed* |
| Treatment duration | Number of years | $\tau$ | 0.5 | Fixed | No | No | (12,13) |
| Rate of on-treatment mortality | Per person per year | $\mu T_j = \frac{k_j}{\tau}$ | Country-specific | Varying | Yes; value for children **greater** than value for adults. | Yes | (14) |
| Rate of treatment completion | Per person per year | $\frac{s_j}{\tau}$ | Country-specific | Fixed equation | Yes, indirectly scaled by $k_{mort}$ | Yes | (14) |
| Rate of treatment non-completion | Per person per year | $\frac{f_j}{\tau}$ | Country-specific | Fixed equation | Yes, indirectly scaled by $k_{mort}$ | Yes | (14) |

### 2.2 Age varying parameters

We assume that aspects of TB natural history and mortality vary by age. This is implemented by stratifying certain natural history parameters by age and applying age-specific prior ranges and relative constraints during calibration (15). The following table describes the method used to operationalise the age varying differences in parameters between adults (ages ≥15 years) and children (ages <15 years). For the rates per year of progression to TB disease, we assumed that the rate for children is less than that for adults. For mortality rates, we assumed the opposite: the rate for children is higher than that for adults.

**Table S 2 How age varying parameters are operationalized.**

| Parameter | Range | Age Varying Description | Age Scaling Parameter | Adults (age 15+) | Children (age 0–14) |
|---|---|---|---|---|---|
| $nonsub_j$ Rate per year of fast progression from Infection to asymptomatic TB | (0.01–0.1) | Retain if value for children is less than value for adults | Sample $k_{prog}$ from (0.5–1) | Sample $infsub_{A15}$ from (0.01–0.1) | $\max(0.01, infsub_{A15} \times k_{mort})$ |
| $subclin_j$ Rate per year progression from asymptomatic TB disease to symptomatic disease | (0.56–0.94) | Retain if value for children is less than value for adults | Sample $k_{prog}$ from (0.5–1) | Sample $infsub_{A15}$ from (0.56–0.94) | max(0.56, $infsub_{A15}$ x $k_{prog}$) |
| $\eta_j$ Rate per year of treatment initiation | (0–2) | Retain if value for children is less than value for adults | Sample $k_{dx}$ from (0.5–1) | Sample $\eta_{A15}$ from (0–2) | max(0, $\eta_{A15}$ x $k_{dx}$) |
| $\mu TB_j$ Symptomatic TB mortality rate per year | (0.28–0.38) | Retain if value for children is greater than value for adults | Sample $k_{mort}$ from (1–1.5) | $\mu TB_{A0}$ x $k_{mort}$ | Sample $\mu TB_{A0}$ from (0.28–0.38) |
| $\mu T_j = \frac{k_j}{\tau}$ On-treatment mortality rate per year | $\left(0 - \frac{k_{max}}{\tau}\right)$ | Retain if value for children is greater than value for adults | Sample $k\_mort$ from (1–1.5) | $\frac{K_{A0}}{\tau} \times k_{mort}$ | Sample $\kappa_{A0}$ from $(0\text{–}0.135)$ |

### 2.3 Treatment initiation and outcomes

Steps for calculating treatment initiation, treatment completion, non-completion, and mortality rates are described in section 3 of the Supplementary Material for Clark et al., *Lancet Glob Health*, 2023 and summarised in Table S 3 below (16).

**Table S 3 Calculating treatment outcome parameter values for adults and children**

| Parameter | Adults | Children |
|---|---|---|
| $\kappa_j$<br>On-treatment mortality fraction | $\kappa_{A0} \times \kappa_{mort}$ | Sample $\kappa_{A0}$ from $(0, 0.135)$ |
| $s_j$<br>On-treatment completion fraction | $(1 - \kappa_{A15})SFR$ | $(1 - \kappa_{A0})SFR$ |
| $f_j$<br>On-treatment non-completion fraction | $(1 - \kappa_{A15})(1 - SFR)$ | $(1 - \kappa_{A0})(1 - SFR)$ |

SFR is the ratio between treatment completions to the sum of the number of treatment completions and non-completions. In India, SFR = 0.96. The data used to calculate the on-treatment outcomes was obtained from the WHO. However, as the private sector accounts for a substantial portion of treatments in India, and not all the treatments conducted in the private sector are reported, we adjust the on-treatment completion and non-completion fractions from Table S 3 as described below and in Table S 4. Additionally, each of the parameters in Table S 3 were divided by $\tau$ to obtain the on-treatment mortality rate per year, on-treatment completion rate per year, and on-treatment non-completion rate per year.

As described in Clark et al., *BMC Medicine*, 2023 (included here with minor text modifications), we adjusted the treatment outcomes to account for treatment occurring in both the public and private sectors (1). We assumed that the total number of treatments was composed of the treatments that are reported and the treatments that are not reported. We assumed that 60% of the total treatment occurs in the public sector and the remaining 40% occurred in the private sector. We assumed that all treatments not reported were from the private sector, that the treatment completion rate in the private sector was 40%, and that there was no reporting bias (in that they were equally likely to not report treatment completions, non-completions, or deaths). Before 2012, only the treatment conducted in the public sector was reported, but since then, treatment in the private sector has begun to be reported, which is reflected by the increasing total fraction of treatments reported (17). We assumed that the on-treatment mortality fraction was the same in the public and private sector but adjusted the treatment completion and non-completion rates to account for differences between those reported and those not reported as in Table S 4.

**Table S 4 Calculation of treatment outcomes for India by year**

| Description | Symbol | Year ($\boldsymbol{k}$) | | | | | | | | |
|---|---|---|---|---|---|---|---|---|---|---|
| | | **≤2012** | **2013** | **2014** | **2015** | **2016** | **2017** | **2018** | **2019** | **≥2020** |
| Fraction of total treatments reported | $F_{T,k}$ | 0.60 | 0.63 | 0.68 | 0.67 | 0.73 | 0.77 | 0.80 | 0.83 | 0.87 |
| On-treatment mortality rate | $\frac{\kappa_j}{\tau}$ | Sample $\kappa_{A0}$ from $(0, 0.135)$ then $\kappa_{A15} = \kappa_{A0} \times \kappa_{mort}$ | | | | | | | | |
| On-treatment completion rate | $\frac{s_j}{\tau}$ | $\frac{F_{T,k}0.96(1-\kappa_j) + (1-F_{T,k})0.40}{\tau}$ | | | | | | | | |
| On-treatment non-completion rate | $\frac{f_j}{\tau}$ | $\frac{F_{T,k}0.04(1-\kappa_j) + (1-F_{T,k})(0.60-\kappa_j)}{\tau}$ | | | | | | | | |

### 3. Model simulation and calibration

#### 3.1 Model simulation

We specified a system of ordinary differential equations defining the derivatives with respect to time of a set of state variables, to simulate the country-specific TB epidemic between 1900 and 2050. We initialised the simulation by distributing the population between the TB natural history states using a fitted parameter representing the proportion of the population uninfected at the start of the simulation. For each year of the simulation (1900–2050), our model is designed to exactly match the age and country-specific UN population estimates and projections. (5)

#### 3.2 Model calibration

For this India modelling analysis, we followed the same modelling approach as in Clark et al., *BMC Medicine*, 2023 (1).

Broadly, this was as follows:

1. Construct a mechanistic model
2. Calibrate the model by identifying areas of the input parameter space where the output of the mechanistic model was consistent with the historical epidemiologic data
3. Use the calibrated model to simulate and predict future TB epidemiology and new vaccines

In the context of this analysis, step 1 was achieved by creating the compartment differential equation model as specified in Section 1. For step 2, we independently calibrated a model by identifying areas of the parameter space that made the output of the model match the corresponding calibration targets (from Table S 5 below). The model was fitted to the calibration targets using history matching with emulation, a method that allows us to explore high-dimensional parameter spaces efficiently and robustly **(18–20)**. History matching progresses as a series of iterations, called waves, where implausible areas of the parameter space, i.e., areas that are unable to give a match between the model output (e.g., the predicted incidence rate by the model) and the empirical data (e.g., the incidence rate calibration target from the WHO data), are found and discarded. In order to identify implausible parameter sets, emulators, which are statistical approximations of model outputs that are built using a modest number of model runs, are used. Emulators provide an estimate of the value of the model at any parameter set of interest, with the advantage that they are orders of magnitude faster than the model.

History matching with emulation, implemented through the *hmer* package in R (21), considerably reduced the size of the parameter space to investigate. Rejection sampling was then performed on the reduced space to identify at least 500 parameter sets that matched all targets. Once we had obtained 500 parameter sets that produced output consistent with the calibration targets, we used those parameter sets with the mechanistic model to simulate the future (step 3).

Table S 5 shows the calibration targets for India. Modifications to calibration targets were made as described in the supplementary material (section 3.4) of Clark et al., *BMC Medicine*, 2023 (1). Targets for all ages were the TB incidence rate in 2000 and 2020 (289 [99, 578] and 188 [129, 257] per 100,000 population), mortality rate in 2000 and 2020 (67 [57, 79] and 37 [34, 40] per 100,000 population), case notification rate in 2000 and 2020 (177 [142, 212] and 136 [109, 163] per 100,000 population), infectious TB prevalence in 2015 and 2021 (315 [210, 529] and 312 [218, 406] per 100,000), and aTB prevalence ratio in 2020 (0.504 [0.361, 0.797]) (5,7,8,22–26). Targets for children were the incidence rate and case notification rate in 2020 (91 [56, 126] and 33 [26, 40] per 100,000 population, respectively) (5,8,25). Targets for adults were the TB incidence rate and case notification rate in 2020 (224 [138, 310] and 173 [138, 208] per 100,000 population) and infectious TB prevalence in 2021 (394 [276, 512] per 100,000 population) (5,8,23,25).

**Table S 5 India national model calibration targets**

<table>
<tr><th>Calibration Targets</th><th>Year</th><th>Age (years)</th><th>Estimate</th><th>Lower</th><th>Upper</th></tr>
<tr><td rowspan="4">TB incidence rate<br>(per 100,000 population/year)</td><td>2000(7)</td><td>All</td><td>289</td><td>99</td><td>578</td></tr>
<tr><td rowspan="3">2020(25)</td><td>All</td><td>188</td><td>129</td><td>257</td></tr>
<tr><td>0–14</td><td>91</td><td>56</td><td>126</td></tr>
<tr><td>≥15</td><td>224</td><td>138</td><td>310</td></tr>
<tr><td rowspan="2">TB mortality rate<br>(per 100,000 population/year)</td><td>2000(7)</td><td>All</td><td>67</td><td>57</td><td>79</td></tr>
<tr><td>2020(7)</td><td>All</td><td>37</td><td>34</td><td>40</td></tr>
<tr><td rowspan="4">TB case notification rate<br>(per 100,000 population/year)</td><td>2000(5,8)</td><td>All</td><td>177</td><td>142</td><td>212</td></tr>
<tr><td rowspan="3">2020(5,8)</td><td>All</td><td>136</td><td>109</td><td>163</td></tr>
<tr><td>0–14</td><td>33</td><td>26</td><td>40</td></tr>
<tr><td>≥15</td><td>173</td><td>138</td><td>208</td></tr>
<tr><td rowspan="3">Infectious TB prevalence<br>(per 100,000 population)</td><td>2015(24,26)</td><td>All</td><td>315</td><td>210</td><td>529</td></tr>
<tr><td>2021(23)</td><td>All</td><td>312</td><td>218</td><td>406</td></tr>
<tr><td>2021(23)</td><td>≥15</td><td>394</td><td>276</td><td>512</td></tr>
<tr><td>Asymptomatic-to-symptomatic TB prevalence ratio</td><td>2020(22)</td><td>All</td><td>0.504</td><td>0.361</td><td>0.797</td></tr>
</table>

### 4. Policy scenarios

#### 4.1 No-new-vaccine scenario

The no-new-vaccine scenario assumed non-vaccine TB interventions continued at current levels into the future. As reported country-level data includes the high coverage levels of neonatal BCG vaccination, this was not explicitly modelled, and we assumed that BCG vaccination would not be discontinued over the model time horizon. We assumed the infectiousness of $aTB$ relative to $sTB$ in the no-new-vaccine scenario ranged between $(0.61, 1)$. (9)

#### 4.2 Vaccine scenarios

***Vaccine eligible population***

We assumed that there was no pre-vaccination infection testing. Therefore, even if a vaccine was only effective when delivered to those with current infection at the time of vaccination, we assumed that all individuals would receive the vaccine, and only those with current infection would receive protection.

***Vaccine efficacy and protection from repeat vaccinations***

Aligning with M72/AS01$_E$ Phase 2b trial results which demonstrated 49.7% (2.1–74.2) efficacy to prevent disease in latently infected adults (27), we assumed 50% prevention of disease efficacy. We assumed that protection increases if a subsequent vaccine course is administered while the individual is currently protected by (1 – current protection) times vaccine efficacy. The number of vaccine courses refers to the number of vaccine courses that the individual is currently protected by, not that they have ever received or have ever been protected by.

***Constant vaccine delivery characteristics***

We assumed that all vaccines had 10-years duration of protection on average with exponential waning. All vaccine scenarios were delivered through routine vaccination of 15-year-olds (80% coverage) starting in 2030, a campaign for ages 16–44 in 2030 (scaled up to 70% coverage over five years) and 2040 (reaching 70% coverage over one year).

***Varied vaccine characteristics***

The following vaccine characteristics were varied in this study: Host infection status at the time of vaccination required for vaccine efficacy, mechanism of prevention of disease action, and the relative aTB infectiousness. Table S 6 below outlines the key varied vaccine characteristics for the vaccine scenarios used in the main and sensitivity analyses.

**Table S 6 Varying vaccine characteristics in the main and sensitivity analyses**

| | Characteristic | | | |
|---|---|---|---|---|
| | **Host stages at the time of vaccination required for vaccine efficacy** | **Mechanism of prevention of disease action** | **Waning conditions** | **Relative aTB infectiousness** |
| ***Main analyses*** | | | | |
| Prevention of only infectious symptomatic disease | AI excluding disease | Reduces progression from aTB to sTB via *subclin* | Efficacy lost once individuals progress into sTB | 0.61–1 |
| Prevention of any infectious disease | AI excluding disease | Reduces progression to aTB from I and nTB via *nonsub*, *infsub* and *relsub*. | Efficacy lost once individuals progress into aTB | 0.61–1 |
| Prevention of any disease | AI excluding disease | Reduces progression from infection to nTB/aTB via *infnon*, *infsub*, *relnon*, and *relsub*. | Efficacy lost once individuals progress into nTB/aTB | 0.61–1 |
| ***Sensitivity analyses*** | | | | |
| Prevention of only infectious symptomatic disease | CI excluding disease | Reduces progression from aTB to sTB via *subclin* | Efficacy lost once individuals progress into sTB | 0.61–1 |
| Prevention of any infectious disease | CI excluding disease | Reduces progression to aTB from I and nTB via *nonsub*, *infsub* and *relsub*. | Efficacy lost once individuals progress into aTB | 0.61–1 |
| Prevention of any disease | CI excluding disease | Reduces progression from infection to nTB/aTB via *infnon*, *infsub*, *relnon*, and *relsub*. | Efficacy lost once individuals progress into nTB/aTB | 0.61–1 |
| Prevention of only infectious symptomatic disease | AI excluding disease | Reduces progression from aTB to sTB via *subclin* | Efficacy lost once individuals progress into sTB | Low = 0.61–0.74<br>Medium = 0.74–0.87<br>High = 0.87–1<br>Zero: 0 |
| Prevention of any infectious disease | AI excluding disease | Reduces progression to aTB from I and nTB via *nonsub*, *infsub* and *relsub*. | Efficacy lost once individuals progress into aTB | Low = 0.61–0.74<br>Medium = 0.74–0.87<br>High = 0.87–1<br>Zero: 0 |
| Prevention of any disease | AI excluding disease | Reduces progression from infection to nTB/aTB via *infnon*, *infsub*, *relnon*, and *relsub*. | Efficacy lost once individuals progress into nTB/aTB | Low = 0.61–0.74<br>Medium = 0.74–0.87<br>High = 0.87–1<br>Zero: 0 |
| Prevention of only infectious symptomatic disease | AI including disease | Reduces progression from aTB to sTB via *subclin* | Efficacy lost once individuals progress into sTB | 0.61–1 |
| Prevention of any infectious disease | AI including disease | Reduces progression to aTB from I and nTB via *nonsub*, *infsub* and *relsub*. | Efficacy lost once individuals progress into aTB | 0.61–1 |
| Prevention of any disease | AI including disease | Reduces progression from infection to nTB/aTB via *infnon*, *infsub*, *relnon*, and *relsub*. | Efficacy lost once individuals progress into nTB/aTB | 0.61–1 |
| Prevention of only infectious symptomatic disease | CI including disease | Reduces progression from aTB to sTB via *subclin* | Efficacy lost once individuals progress into sTB | 0.61–1 |
| Prevention of any infectious disease | CI including disease | Reduces progression to aTB from I and nTB via *nonsub*, *infsub* and *relsub*. | Efficacy lost once individuals progress into aTB | 0.61–1 |
| Prevention of any disease | CI including disease | Reduces progression from infection to nTB/aTB via *infnon*, *infsub*, *relnon*, and *relsub*. | Efficacy lost once individuals progress into nTB/aTB | 0.61–1 |

***Vaccine model structure***

The vaccine model structure used in this work was described in section 4.3 of the Supplementary Material of Clark et al., *BMC Medicine*, 2023 (1), and reproduced here with modifications.

The vaccine structure for an any infection (AI) vaccine or a current infection (CI) vaccine is in Figure S 2. Each compartment in the vaccine structure is replicated for all TB natural history compartments and ages. An AI vaccine was assumed to be efficacious with any infection status (aside from current active disease) at the time of vaccination, whereas the CI vaccine was assumed to be efficacious only with current infection at the time of vaccination. We accounted for differences in *Vaccinated Protected*, *Vaccinated Not Protected*, and *Vaccinated Waned*. The *Vaccinated Not Protected* compartments are included as we assume that individuals with may be accidentally vaccinated and would not receive protection from the vaccine. With each vaccine course the level of protection builds if the recipient is currently in a *Vaccinated Protected* compartment. Waning occurs from any of the *Vaccinated Protected* compartments to the *Vaccinated Protected* compartment one level below, or to the *Waned Protection* compartment for those with only one course of protection.

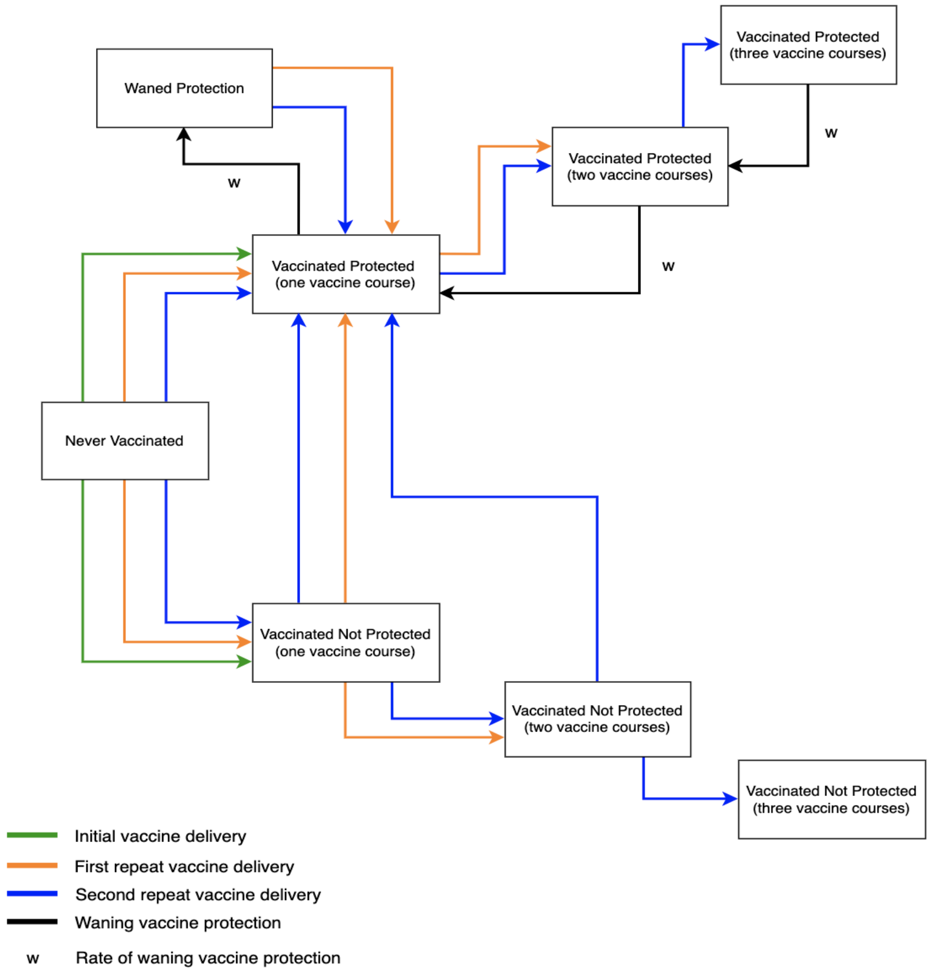


**Figure S 2 Vaccine structure for an AI or CI vaccine**

***Vaccine integration in the TB natural history model***

Vaccine protection is incorporated in the TB natural history structure as indicated with the dashed arrows in Figure S 3. The vaccine acts by reducing the rate of progression to disease by $(1 - pV)$ depending on vaccine scenario characteristics, where $pV$ is the vaccine efficacy.

For the vaccine preventing progression to infectious symptomatic disease only (the red dashed line), we assumed that $(1 - pV)$ would be applied to $subclin$.

For the vaccine preventing progression to any infectious disease (the blue dotted-dashed lines), we assumed that $(1 - pV)$ would be applied to $infsub$, $nonsub$, and $relsub$.

For the vaccine preventing progression to any disease (the grey dotted lines), we assumed that $(1 - pV)$ would be applied to $infnon$, $infsub$, $relnon$, and $relsub$.

Vaccine efficacy was modelled as "degree", which assumes that everyone who has been vaccinated and receives protection (those in the *Vaccinated Protected* compartments) will have protection from the vaccine equivalent to the value of the vaccine efficacy.

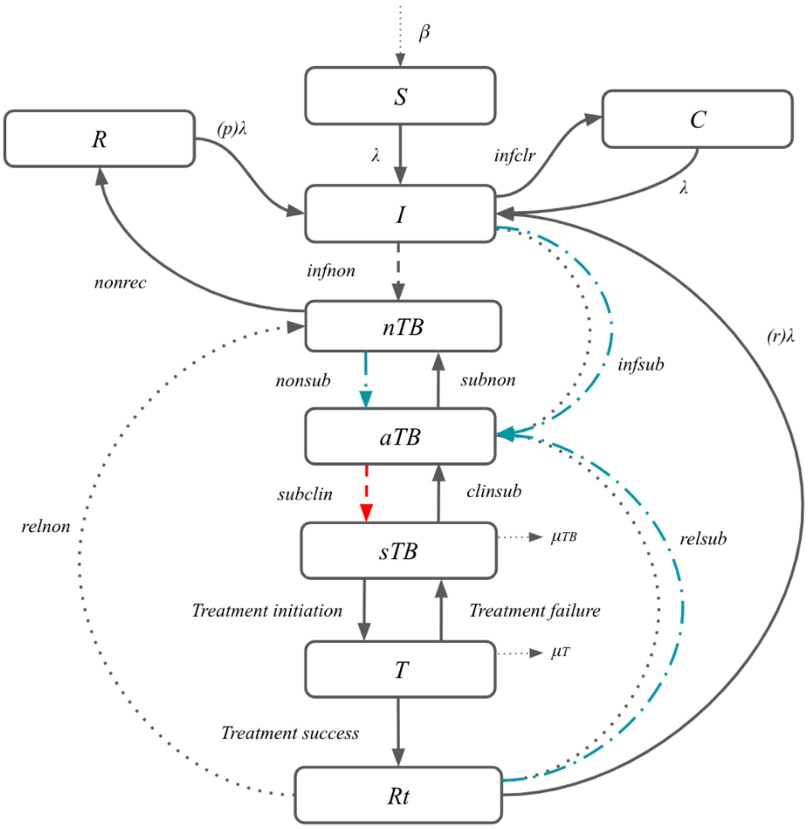


**Figure S 3** **TB natural history structure indicating where vaccine efficacy is applied**

The host infection status required for vaccine efficacy is indicated with the shaded compartments in Figure S 4. For scenarios where we assumed that all vaccine mechanisms of action (preventing progression to only infectious symptomatic disease, any infectious disease, or any disease) would not be effective in pre-symptomatic stages, we assumed:

- AI – excluding disease: grey shaded compartments (both solid and lined)
- CI – excluding disease: grey shaded compartments (with lines only)

For scenarios where we assumed the vaccine would be effective in pre-symptomatic stages, we assumed that for the vaccine preventing progression to symptomatic disease only:

- AI – including disease: grey shaded compartments (both solid and lined), the blue compartment, and the red compartment
- CI – including disease: grey shaded compartments (with lines only), the blue compartment, and the red compartment

For the vaccine preventing progression to any infectious disease:

- AI – including disease: grey shaded compartments (both solid and lined) and the blue compartment
- CI – including disease: grey shaded compartments (with lines only) and the blue compartment

There was no scenario including disease for a vaccine preventing progression to any disease, as there were no pre-symptomatic stages before where the vaccine would exert the prevention of disease effect.

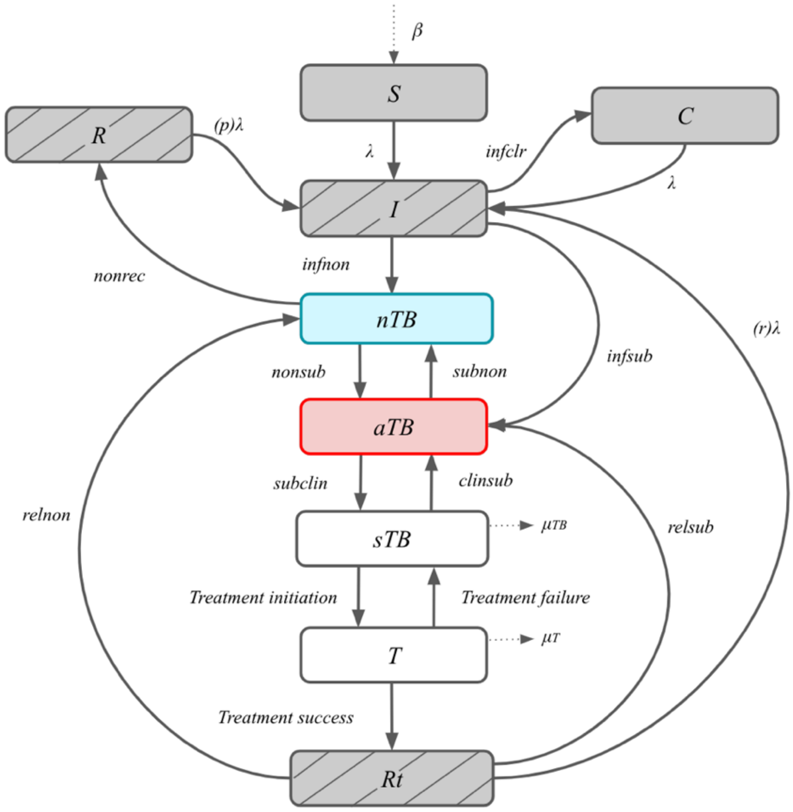


**Figure S 4** **TB natural history structure indicating host infection status required for efficacy**

### SUPPORTING RESULTS

### 5. No-new-vaccine scenario results

#### 5.1 No-new-vaccine calibration (with baseline, low, medium and high relative infectiousness)

Figure S 5 shows trends in TB incidence, TB case notifications, infectious TB disease prevalence, TB mortality, proportion of infectious TB that is asymptomatic, and TB infection prevalence from 2000-2050 for all ages, based on 500 parameter sets calibrated to all 14 targets under varying assumptions about relative asymptomatic TB infectiousness: baseline (0.61–1), low (0.61–0.74), medium (0.74–0.87), and high (0.87–1).

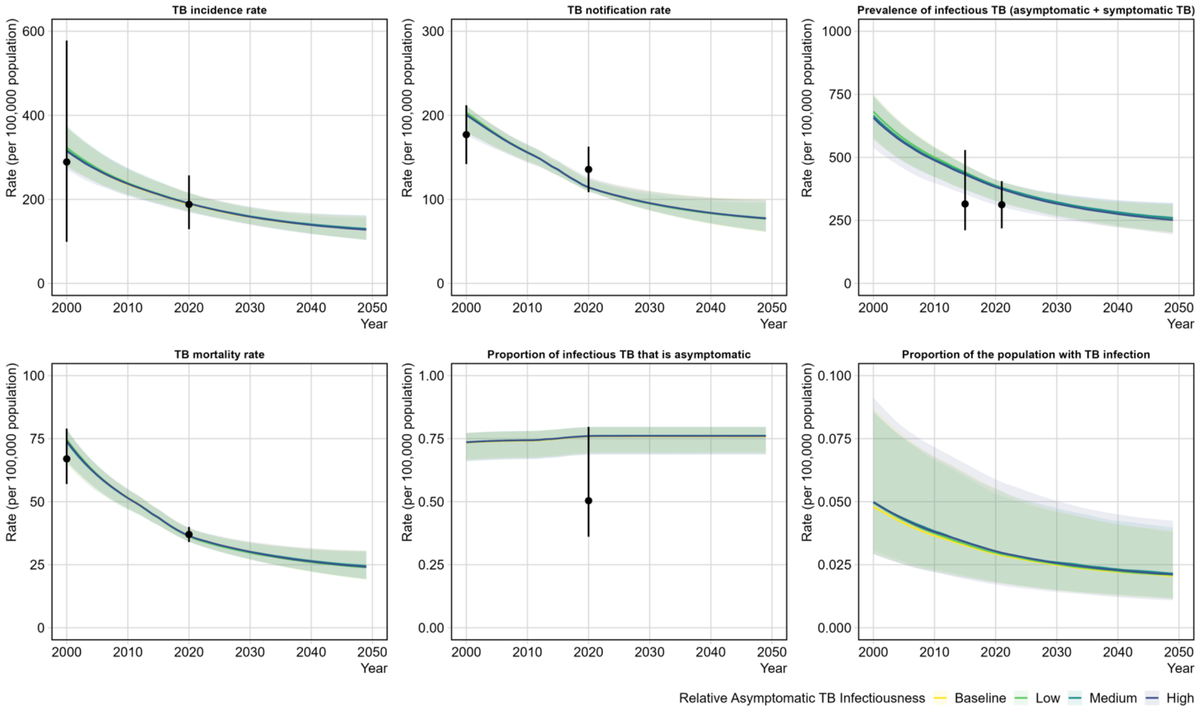


**Figure S 5 Trends in TB epidemiology from 2000–2050 for all ages for model calibrations under varying assumptions about relative asymptomatic TB infectiousness: baseline (0.61–1), low (0.61–0.74), medium (0.74–0.87), and high (0.87–1).** The trend lines in yellow, green, blue and purple indicate the median modelled output with 95% uncertainty reflected by the respective shaded colours. The black dot and vertical lines are the calibration targets from Table S 5. Note y-axis scales differ.

### 5.2 No-new-vaccine scenario calibration (with zero relative infectiousness)

Figure S 6 shows trends in tuberculosis incidence, TB case notifications, infectious TB disease prevalence, TB mortality, proportion of infectious TB that is asymptomatic, and TB infection prevalence from 2000–2050 for all ages, based on 500 parameter sets calibrated to 13 out of 14 calibration targets under zero relative aTB infectiousness. The zero-infectiousness scenario fit to 13 out of 14 targets, missing either TB mortality or the TB case notification target for individuals aged 0–99 in the year 2000 (Figure S 6).

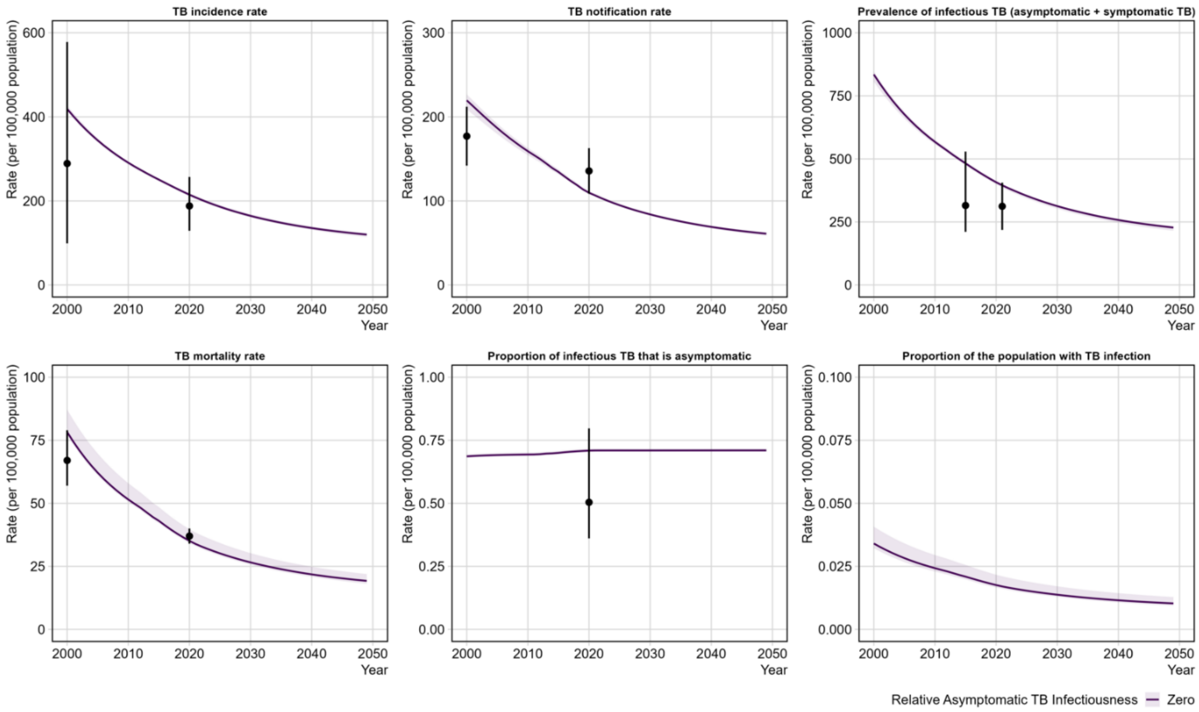


**Figure S 6 Trends in TB epidemiology from 2000–2050 for all ages for zero relative aTB infectiousness.** Note y-axis scales differ.

Table S 7 shows the number of sTB, aTB, and nTB episodes, the overall number of infectious episodes (aTB + sTB), and TB deaths for the no-new-vaccine scenario under the baseline and zero aTB relative infectiousness scenarios.

**Table S 7 No-new-vaccine baseline incidence of TB episodes and deaths (millions), under baseline and zero aTB infectiousness relative to sTB**

| Averted numbers (millions) | Short-term (2030–2032) | | Longer-term (2030–2050) | |
|---|---|---|---|---|
| | Baseline infectiousness (0.61–1) | Zero infectiousness (0) | Baseline infectiousness (0.61–1) | Zero infectiousness (0) |
| sTB | 7.10 (6.33, 8.14) | 7.33 (7.17, 7.54) | 47.04 (40.45, 56.24) | 46.10 (44.63, 47.48) |
| aTB | 26.72 (21.17, 31.29) | 27.08 (26.30, 27.64) | 175.84 (136.35, 216.28) | 170.35 (163.82, 174.56) |
| aTB + sTB | 33.82 (27.93, 39.05) | 34.51 (33.48, 35.04) | 222.81 (178.48, 272.62) | 217.08 (208.84, 221.19) |
| nTB | 35.58 (25.74, 47.09) | 33.55 (31.92, 36.39) | 233.91 (166.48, 324.57) | 211.95 (199.59, 230.41) |
| TB deaths | 1.34 (1.22, 1.53) | 1.18 (1.14, 1.35) | 8.84 (7.51, 10.65) | 7.43 (7.11, 8.46) |

*Abbreviations: aTB = infectious asymptomatic TB; nTB = non-infectious TB; sTB = infectious symptomatic TB*

### 6. Vaccinated proportions over time

Figure S 7 shows the annual vaccinated proportions between 2030 and 2050 for ages 15, 25 and 40-year-olds across the *Basecase* scenarios.

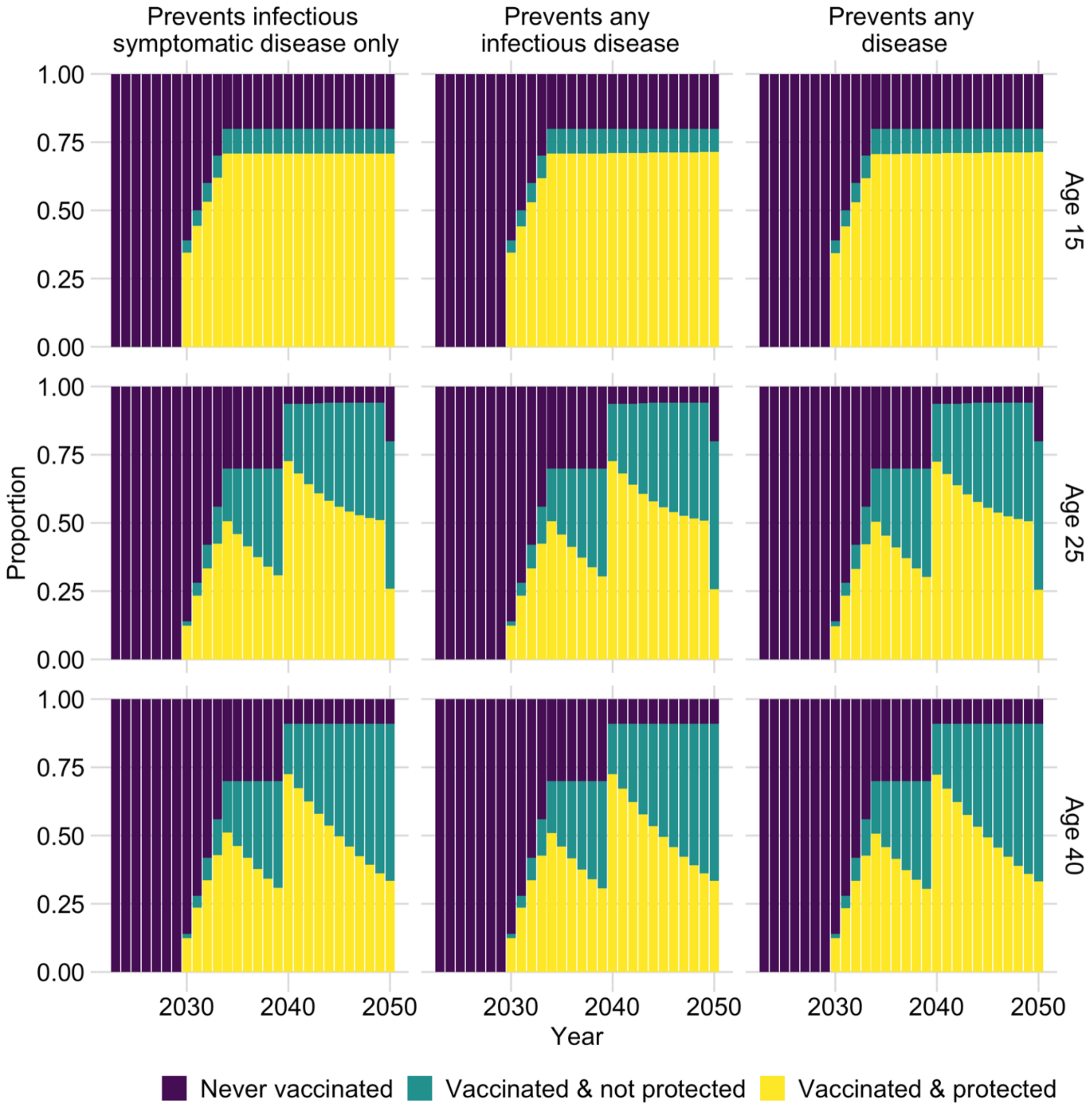


**Figure S 7 Proportion of population vaccinated from 2030–2050, for ages 15, 25 and 40 years old.**

### 7. Sensitivity analysis results: vaccines effective with current infection status

Figure S 8 and Table S 8 shows the short-term and longer-term impact of vaccines that are effective in current infection status at the time of vaccination. Results showed that vaccines effective only in infected individuals had lower impact at all time points compared to the *Basecase* analysis but displayed similar patterns and trends in their mechanism of action to vaccines effective in both uninfected and infected individuals.

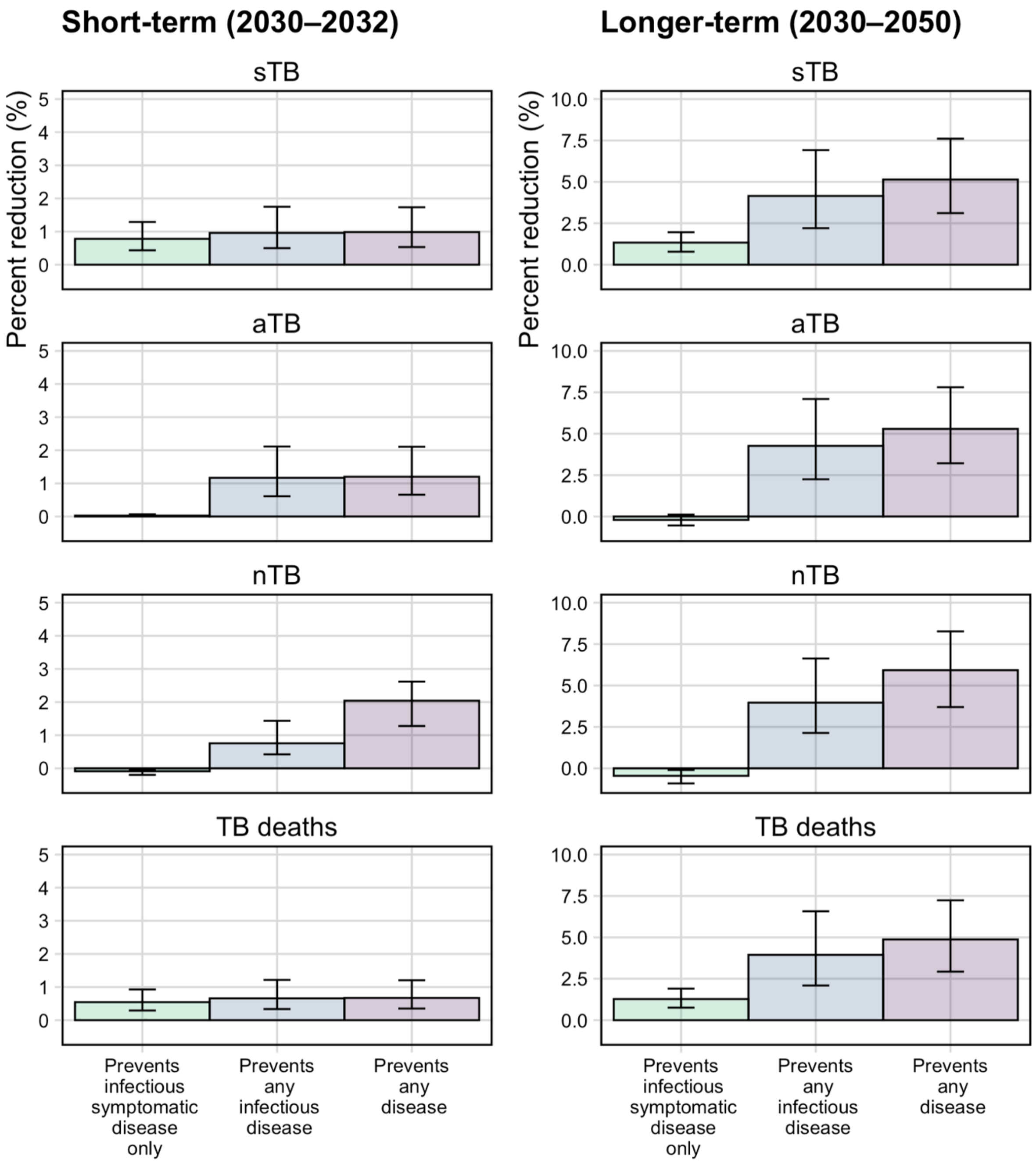


**Figure S 8 Percentage cumulative TB episodes and deaths averted between 2030–2023 (left column) or between 2030–2050 (right column) for a current infection vaccine.** Note y-axis scales differ. *Abbreviations: aTB = infectious asymptomatic TB; nTB = non-infectious TB; sTB = infectious symptomatic TB.*

**Table S 8 Number and percentage cumulative TB episodes and deaths averted (2030–2032 and 2030–2050) for vaccines effective with current infection under baseline aTB infectiousness**

| | | 2030–2032 | | | 2030–2050 | | |
|---|---|---|---|---|---|---|---|
| **Vaccine prevents progression to:** | | **Infectious symptomatic disease only** | **Any infectious disease** | **Any disease** | **Infectious symptomatic disease only** | **Any infectious disease** | **Any disease** |
| **Episodes averted (millions)** | sTB | 0.06 (0.03, 0.10) | 0.07 (0.03, 0.13) | 0.07 (0.04, 0.12) | 0.63 (0.34, 1.01) | 1.97 (0.97, 3.48) | 2.43 (1.37, 3.85) |
| | aTB | 0.01 (0.00, 0.02) | 0.31 (0.16, 0.56) | 0.32 (0.17, 0.56) | -0.37 (-0.94, 0.21) | 7.45 (3.71, 13.23) | 9.28 (5.25, 15.07) |
| | aTB + sTB | 0.06 (0.03, 0.11) | 0.38 (0.20, 0.69) | 0.39 (0.21, 0.69) | 0.21 (-0.35, 1.08) | 9.43 (4.75, 16.54) | 11.65 (6.65, 18.89) |
| | nTB | -0.03 (-0.06, -0.01) | 0.28 (0.15, 0.45) | 0.70 (0.41, 1.10) | -1.07 (-2.14, -0.25) | 9.37 (4.72, 15.80) | 13.91 (7.46, 22.53) |
| | TB deaths | 0.01 (0.00, 0.01) | 0.01 (0.00, 0.02) | 0.01 (0.00, 0.02) | 0.11 (0.06, 0.19) | 0.35 (0.18, 0.62) | 0.44 (0.24, 0.69) |
| **Percent reduction (%)** | sTB | 0.78 (0.44, 1.29) | 0.96 (0.50, 1.75) | 0.98 (0.53, 1.73) | 1.33 (0.78, 1.96) | 4.15 (2.20, 6.92) | 5.15 (3.11, 7.61) |
| | aTB | 0.03 (0.01, 0.07) | 1.17 (0.61, 2.11) | 1.20 (0.66, 2.11) | -0.20, (-0.54, 0.12) | 4.27 (2.25, 7.10) | 5.29 (3.22, 7.81) |
| | aTB + sTB | 0.19 (0.10, 0.33) | 1.12 (0.59, 2.04) | 1.16 (0.63, 2.03) | 0.10 (-0.16, 0.47) | 4.24 (2.24, 7.05) | 5.25 (3.19, 7.76) |
| | nTB | -0.09 (-0.20, -0.04) | 0.76 (0.42, 1.43) | 2.04 (1.28, 2.62) | -0.46 (-0.91, -0.10) | 3.97 (2.14, 6.63) | 5.93 (3.70, 8.27) |
| | TB deaths | 0.54 (0.29, 0.93) | 0.66 (0.33, 1.22) | 0.67 (0.35, 1.20) | 1.28 (0.75, 1.90) | 3.94 (2.09, 6.58) | 4.87 (2.93, 7.24) |

*Abbreviations: aTB = infectious asymptomatic TB; nTB = non-infectious TB; sTB = infectious symptomatic TB*

### 8. Trends over time for vaccines effective with current infection status

Figure S 9 shows the trends in the number of sTB, aTB, and nTB episodes and TB deaths between 2030–2050 for each current infection vaccine scenario compared to no-new-vaccine scenario, under baseline relative aTB infectiousness (0.61–1).

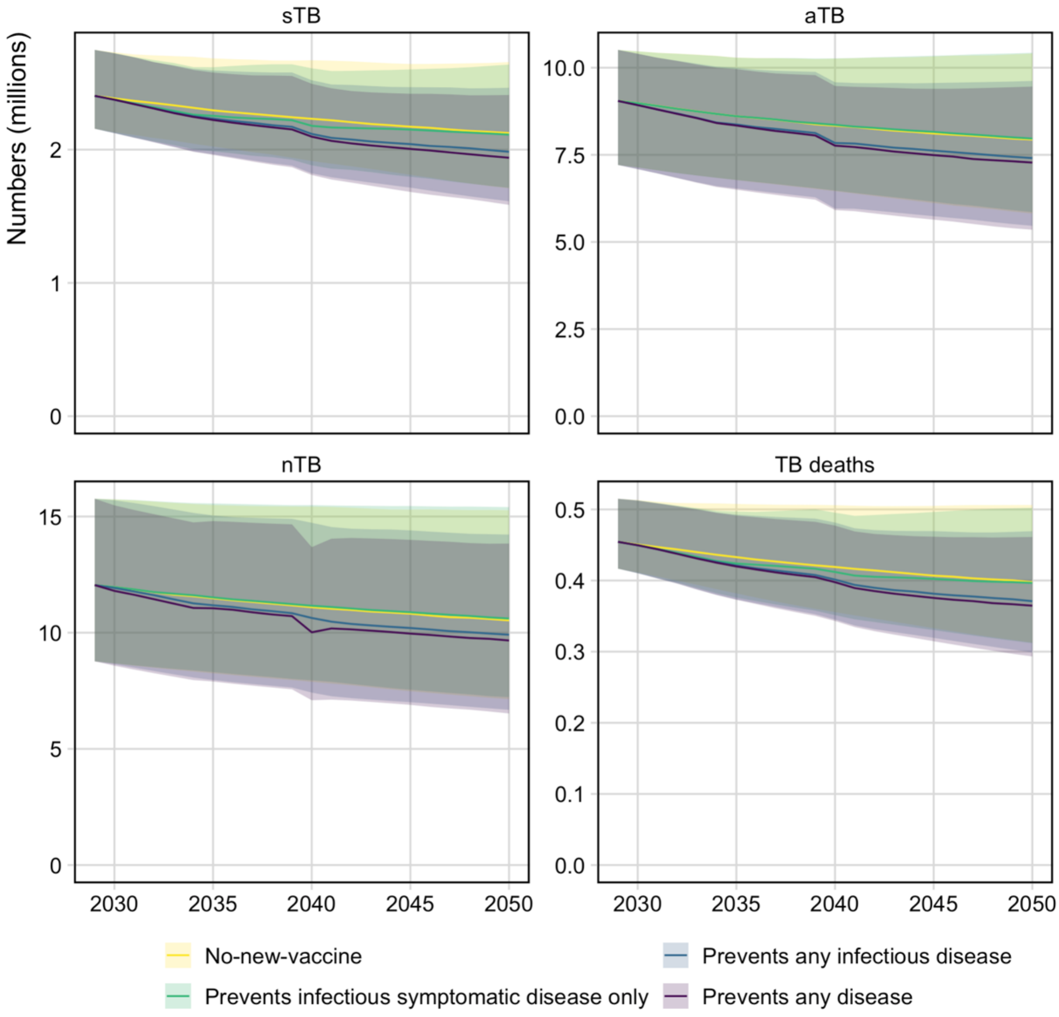


**Figure S 9 Trends in the number of sTB, aTB, and nTB episodes and TB deaths between 2030–2050 for each current infection vaccine scenario compared to no-new-vaccine scenario for baseline aTB infectiousness.** Note y-axis scales differ.

### 9. Sensitivity analysis results: vaccines effective with any infection status with varying infectiousness

#### 9.1 Short-term impact (2030–2032)

The short-term impact of vaccines that are effective in any infection status at the time of vaccination under varying assumptions about the relative infectiousness of aTB is in Figure S 10 and

Table S 9. Infectiousness scenarios were based on current literature estimates (0.61–1) and divided into low (0.61–0.74), medium (0.74–0.87), and high (0.87–1) ranges, with an additional scenario assuming zero infectiousness. Results showed that the short-term impact of vaccines across all infectiousness scenarios was similar to the *Basecase* vaccine scenario impact estimates, which assumed baseline infectiousness (0.61–1) (Figure S 10,

Table S 9).

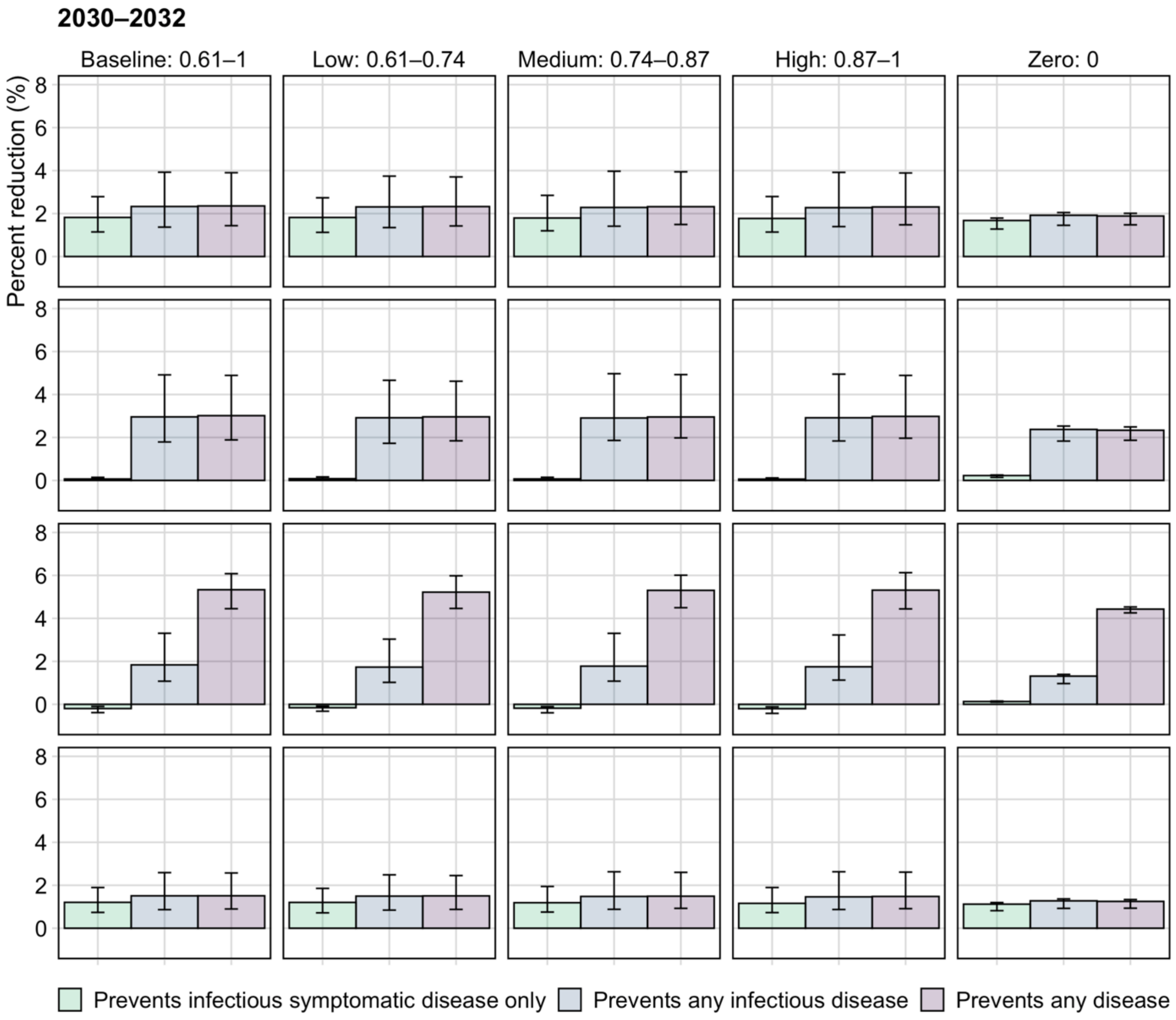


**Figure S 10 Percentage cumulative TB episodes and deaths averted between 2030–2032 for an any infection vaccine by varied relative infectiousness of aTB.**

*Abbreviations: aTB = infectious asymptomatic TB; nTB = Non-infectious TB; sTB = infectious symptomatic TB*

**Table S 9 Number and percentage cumulative TB episodes and deaths averted between 2030–2032 for vaccines effective in any infection, under low, medium, high and zero asymptomatic TB infectiousness relative to symptomatic TB**

| | | Episodes Averted (millions) | | | Percent reduction (%) | | |
|---|---|---|---|---|---|---|---|
| Vaccine prevents progression to: | | Infectious symptomatic disease only | Any infectious disease | Any disease | Infectious symptomatic disease only | Any infectious disease | Any disease |
| **Low infectiousness (0.61-0.74)** | nTB | -0.06 (-0.10, -0.03) | 0.63 (0.35, 1.02) | 1.90 (1.32, 2.60) | -0.16 (-0.32, -0.08) | 1.73 (1.02, 3.03) | 5.22 (4.46, 5.98) |
| | aTB | 0.02 (0.01, 0.05) | 0.77 (0.43, 1.34) | 0.79 (0.46, 1.33) | 0.08 (0.04, 0.16) | 2.92 (1.73, 4.66) | 2.96 (1.84, 4.62) |
| | sTB | 0.13 (0.08, 0.20) | 0.16 (0.09, 0.27) | 0.16 (0.10, 0.27) | 1.82 (1.13, 2.73) | 2.31 (1.35, 3.74) | 2.32 (1.42, 3.71) |
| | aTB + sTB | 0.15 (0.09, 0.24) | 0.94 (0.52, 1.60) | 0.96 (0.55, 1.60) | 0.44 (0.27, 0.68) | 2.79 (1.64, 4.48) | 2.83 (1.75, 4.44) |
| | TB deaths | 0.02 (0.01, 0.03) | 0.02 (0.01, 0.03) | 0.02 (0.01, 0.03) | 1.20 (0.72, 1.85) | 1.49 (0.84, 2.49) | 1.51 (0.88, 2.45) |
| **Medium infectiousness (0.74-0.87)** | nTB | -0.07 (-0.12, -0.04) | 0.65 (0.36, 1.07) | 1.88 (1.30, 2.71) | -0.18% (-0.39, -0.11) | 1.78% (1.08, 3.30) | 5.30% (4.50, 6.01) |
| | aTB | 0.02 (0.01, 0.04) | 0.79 (0.44, 1.42) | 0.80 (0.46, 1.41) | 0.07% (0.03, 0.15) | 2.91% (1.86, 4.97) | 2.95% (1.98, 4.92) |
| | sTB | 0.13 (0.08, 0.21) | 0.16 (0.09, 0.29) | 0.17 (0.10, 0.29) | 1.79% (1.20, 2.85) | 2.28% (1.41, 3.97) | 2.32% (1.49, 3.94) |
| | aTB + sTB | 0.15 (0.09, 0.24) | 0.95 (0.54, 1.71) | 0.97 (0.57, 1.69) | 0.43% (0.27, 0.69) | 2.77% (1.77, 4.76) | 2.82% (1.88, 4.73) |
| | TB deaths | 0.02 (0.01, 0.03) | 0.02 (0.01, 0.04) | 0.02 (0.01, 0.04) | 1.19% (0.75, 1.94) | 1.48% (0.88, 2.63) | 1.49% (0.93, 2.60) |
| **High infectiousness (0.87-1)** | nTB | -0.07 (-0.12, -0.04) | 0.64 (0.34, 1.02) | 1.84 (1.24, 2.72) | -0.20% (-0.42, -0.12) | 1.75% (1.13, 3.23) | 5.31% (4.44, 6.13) |
| | aTB | 0.02 (0.01, 0.03) | 0.76 (0.43, 1.36) | 0.78 (0.46, 1.34) | 0.06% (0.03, 0.12) | 2.92% (1.84, 4.95) | 2.98% (1.96, 4.89) |
| | sTB | 0.13 (0.08, 0.20) | 0.16 (0.10, 0.29) | 0.17 (0.10, 0.28) | 1.77% (1.14, 2.79) | 2.27% (1.39, 3.92) | 2.31% (1.48, 3.89) |
| | aTB + sTB | 0.14 (0.09, 0.23) | 0.93 (0.53, 1.64) | 0.94 (0.56, 1.62) | 0.43% (0.28, 0.65) | 2.77% (1.75, 4.74) | 2.82% (1.86, 4.71) |
| | TB deaths | 0.02 (0.01, 0.03) | 0.02 (0.01, 0.04) | 0.02 (0.01, 0.04) | 1.16% (0.73, 1.90) | 1.46% (0.87, 2.63) | 1.48% (0.91, 2.61) |
| **Zero infectiousness (0)** | nTB | 0.04 (0.04, 0.05) | 0.43 (0.35, 0.47) | 1.48 (1.37, 1.62) | 0.13% (0.10, 0.15) | 1.31% (0.97, 1.39) | 4.43% (4.26, 4.53) |
| | aTB | 0.06 (0.04, 0.07) | 0.64 (0.50, 0.69) | 0.63 (0.51, 0.68) | 0.23% (0.14, 0.25) | 2.37% (1.83, 2.53) | 2.34% (1.87, 2.49) |
| | sTB | 0.12 (0.10, 0.13) | 0.14 (0.11, 0.15) | 0.14 (0.11, 0.15) | 1.68% (1.28, 1.79) | 1.92% (1.45, 2.05) | 1.89% (1.48, 2.01) |
| | aTB + sTB | 0.18 (0.13, 0.20) | 0.78 (0.60, 0.84) | 0.77 (0.62, 0.83) | 0.53% (0.39, 0.58) | 2.28% (1.75, 2.43) | 2.24% (1.78, 2.39) |
| | TB deaths | 0.01 (0.01, 0.01) | 0.02 (0.01, 0.02) | 0.01 (0.01, 0.02) | 1.12% (0.82, 1.20) | 1.27% (0.92, 1.36) | 1.25% (0.93, 1.34) |

*Abbreviations: aTB = infectious asymptomatic TB; nTB = non-infectious TB; sTB = infectious symptomatic TB.*

#### 9.2 Longer-term impact (2030–2050)

Figure S 11 and Table S 10 shows the longer-term impact of vaccines that are effective with any infection status at the time of vaccination under varying assumptions about the relative infectiousness of aTB. Results showed that the longer-term impact of vaccines under the low (0.61–0.74), medium (0.74–0.87), and high (0.87–1) infectiousness scenarios was similar to the *Basecase* vaccine scenario impact estimates, which assumed baseline infectiousness (0.61–1). However, under the zero infectiousness scenario (assuming aTB is not infectious), vaccines that prevented progression to any infectious disease or any disease showed reduced impact across all measured outcomes, whereas the vaccine that prevented progression to only infectious symptomatic disease showed increased impact across all measured outcomes compared to the *Basecase* vaccine scenario impact estimates (
Figure S 11, Table S 10).

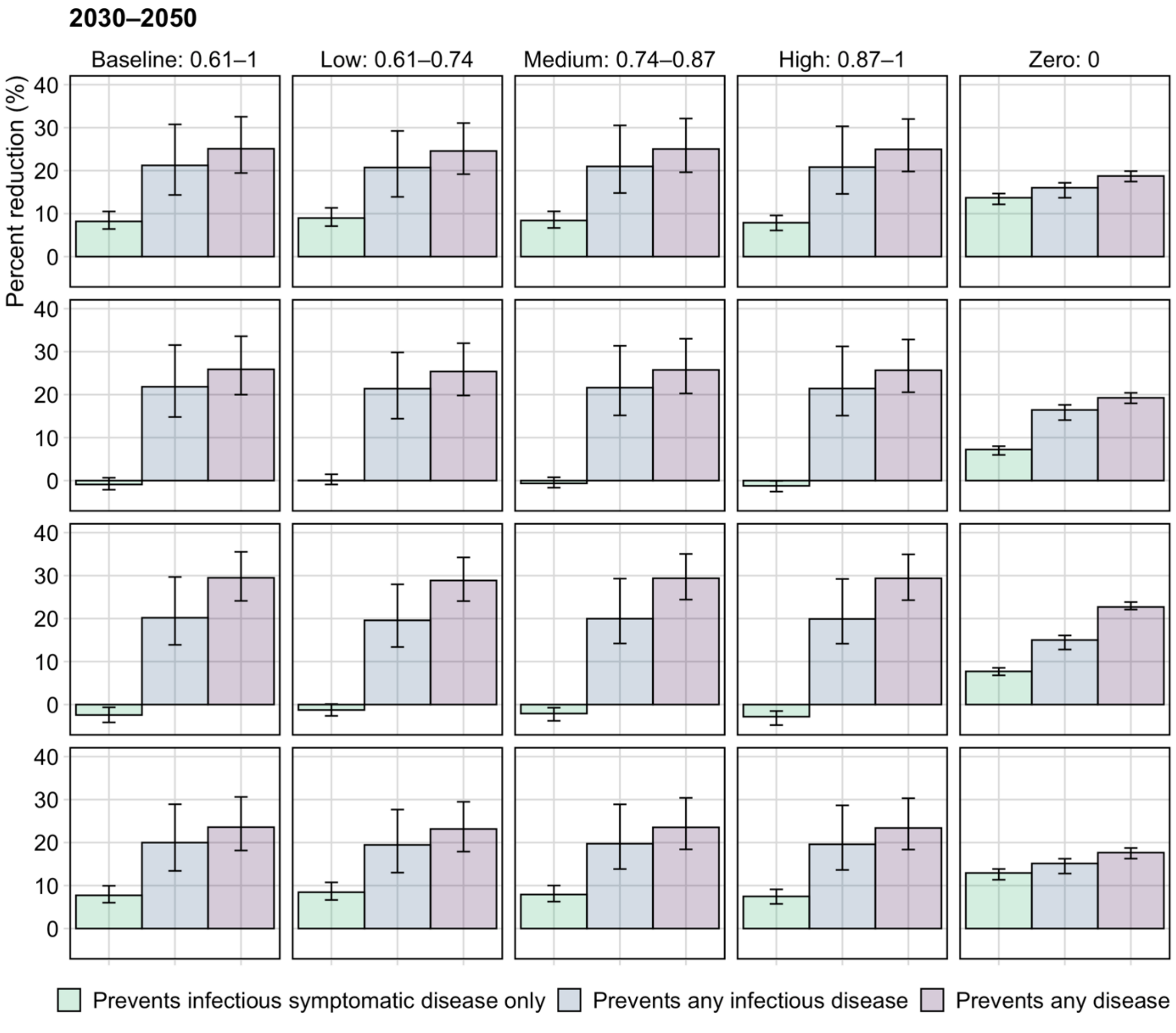


**Figure S 11 Percentage cumulative TB episodes and deaths averted between 2030-2050 for an any infection vaccine by varied relative infectiousness of aTB**

*Abbreviations: aTB = infectious asymptomatic TB; nTB = non-infectious TB; sTB = infectious symptomatic TB*

**Table S 10 Number and percentage cumulative TB episodes and deaths averted between 2030–2050 for vaccines effective in any infection, under low, medium, high and zero asymptomatic TB infectiousness relative to symptomatic TB.**

| | | Numbers averted (millions) | | | Percent reduction (%) | | |
|---|---|---|---|---|---|---|---|
| Vaccine prevents progression to: | | Infectious symptomatic disease only | Any infectious disease | Any disease | Infectious symptomatic disease only | Any infectious disease | Any disease |
| Low infectiousness (0.61–0.74) | sTB | 4.23 (2.97, 5.79) | 9.81 (6.06, 14.47) | 11.69 (8.18, 15.7) | 8.98% (7.09, 11.34) | 20.73% (13.9, 29.23) | 24.58% (19.18, 31.08) |
| | aTB | 0.07 (-1.61, 2.76) | 37.44 (22.3, 59.31) | 44.92 (30.2, 63.95) | 0.04% (-0.91, 1.48) | 21.38% (14.4, 29.82) | 25.37% (19.8, 31.95) |
| | aTB + sTB | 4.25 (1.78, 8.10) | 47.09 (28.19, 73.05) | 56.64 (38.11, 79.68) | 1.89% (0.88, 3.49) | 21.26% (14.28, 29.71) | 25.21% (19.66, 31.78) |
| | nTB | -3.06 (-5.83, 0.29) | 47.42 (28.51, 70.55) | 69.32 (47.20, 98.46) | -1.27% (-2.63, 0.13) | 19.59% (13.39, 27.97) | 28.87% (24.06, 34.23) |
| | TB deaths | 0.74 (0.52, 1.06) | 1.70 (1.04, 2.60) | 2.04 (1.40, 2.85) | 8.44% (6.65, 10.73) | 19.46% (13.01, 27.67) | 23.16% (17.89, 29.48) |
| Medium infectiousness (0.74–0.87) | sTB | 4.01 (2.80, 5.56) | 10.10 (6.20, 15.26) | 12.04 (8.30, 16.22) | 8.40% (6.67, 10.54) | 20.99% (14.79, 30.52) | 25.03% (19.63, 32.12) |
| | aTB | -1.16 (-2.92, 1.45) | 38.71 (22.45, 63.46) | 46.24 (29.42, 67.55) | -0.65% (-1.65, 0.76) | 21.61% (15.17, 31.36) | 25.74% (20.26, 33.00) |
| | aTB + sTB | 2.79 (0.77, 6.79) | 48.66 (29.00, 77.94) | 57.77 (38.45, 82.88) | 1.24% (0.36, 2.90) | 21.47% (15.09, 31.18) | 25.58% (20.14, 32.80) |
| | nTB | -5.07 (-8.23, -1.88) | 48.05 (29.02, 72.93) | 69.72 (47.43, 102.69) | -2.09% (-3.78, -0.73) | 19.97% (14.22, 29.29) | 29.38% (24.42, 35.04) |
| | TB deaths | 0.72 (0.49, 0.98) | 1.77 (1.11, 2.75) | 2.12 (1.45, 2.92) | 7.92% (6.26, 10.00) | 19.73% (13.84, 28.89) | 23.54% (18.42, 30.40) |
| High infectiousness (0.87–1) | sTB | 3.69 (2.55, 5.27) | 9.94 (6.31, 15.19) | 11.93 (8.39, 16.40) | 7.89% (6.09, 9.58) | 20.83% (14.59, 30.31) | 24.95% (19.80, 32.00) |
| | aTB | -2.13 (-4.30, -0.08) | 37.45 (21.10, 59.68) | 45.13 (28.55, 64.96) | -1.22% (-2.56, -0.04) | 21.41% (15.11, 31.22) | 25.67% (20.56, 32.83) |
| | aTB + sTB | 1.53 (-0.87, 4.61) | 47.35 (27.67, 73.87) | 56.87 (37.67, 80.79) | 0.66% (-0.42, 2.00) | 21.28% (14.98, 31.02) | 25.48% (20.38, 32.58) |
| | nTB | -6.58 (-10.06, -3.64) | 47.30 (26.31, 70.69) | 67.91 (43.60, 101.54) | -2.82% (-4.78, -1.51) | 19.91% (14.17, 29.21) | 29.37% (24.28, 34.94) |
| | TB deaths | 0.67 (0.44, 0.93) | 1.76 (1.08, 2.66) | 2.10 (1.43, 2.88) | 7.48% (5.73, 9.12) | 19.60% (13.63, 28.66) | 23.39% (18.37, 30.31) |
| Zero infectiousness (0) | sTB | 6.23 (5.74, 6.85) | 7.29 (6.47, 8.03) | 8.53 (8.19, 9.27) | 13.70% (12.15, 14.67) | 16.02% (13.70, 17.18) | 18.75% (17.46, 19.88) |
| | aTB | 12.21 (10.18, 13.75) | 27.76 (23.96, 30.27) | 32.54 (30.60, 35.11) | 7.22% (5.98, 8.01) | 16.43% (14.07, 17.61) | 19.25% (17.98, 20.42) |
| | aTB + sTB | 18.45 (15.92, 20.56) | 35.06 (30.43, 38.22) | 41.08 (38.84, 44.19) | 8.56% (7.32, 9.43) | 16.34% (13.99, 17.52) | 19.15% (17.87, 20.30) |
| | nTB | 16.07 (14.73, 18.34) | 31.21 (29.19, 34.42) | 48.96 (44.58, 51.58) | 7.71% (6.80, 8.53) | 15.00% (12.80, 16.07) | 22.70% (22.11, 23.84) |
| | TB deaths | 0.96 (0.90, 1.04) | 1.10 (1.05, 1.22) | 1.34 (1.23, 1.41) | 12.93% (11.35, 13.85) | 15.13% (12.79, 16.23) | 17.66% (16.26, 18.72) |

*Abbreviations: nTB = non-infectious TB; aTB = infectious asymptomatic TB; sTB = infectious symptomatic TB*

### 10. Sensitivity analysis results: vaccines effective with any or current infection status including efficacy in pre-disease stages

Figure S 12 and Table S 11 shows the short-term vs. longer-term vaccine impact where an any infection or current infection vaccine is also effective in early disease stages.

If the vaccine preventing progression to any infectious disease was also effective if delivered to those with nTB, and the vaccine preventing progression to infectious symptomatic disease only was also effective if delivered to those with nTB and aTB, we observed differences in impact compared to any infection vaccines that were not effective if given to individuals with nTB and aTB.

The impact of a vaccine preventing any infectious disease effective if given to individuals with nTB increased for all outcomes compared to the vaccine that was ineffective if given to individuals with nTB, in both the short- and longer-term, as a higher proportion of the population would receive protection from the vaccine (Figure S 12, Table S 11). In both the short- and longer-term, this increase resulted in the impact of this vaccine being as much as or greater than the impact of a vaccine preventing progression to any disease for all outcomes (Figure S 12, Table S 11).

The impact of a vaccine preventing only infectious symptomatic disease that was effective if given to individuals with nTB or aTB became more pronounced compared to the vaccine that was ineffective if given to individuals with nTB or aTB, with greater increases in nTB or aTB episodes, and greater reductions in sTB episodes and TB deaths (Figure S 12, Table S 11). In the short-term, the greatest impact on averting sTB episodes and TB deaths was from a vaccine preventing only infectious symptomatic disease effective if given to individuals with nTB or aTB, compared to a vaccine preventing any disease, or preventing any infectious disease effective if given to individuals with nTB. However, in the longer term, the relative impact of the vaccine decreases compared to the other vaccines, due to the build-up of the transmission effect (Figure S 12, Table S 11).

When comparing the impact for any infection vaccines effective if given to individuals with nTB or aTB, to current infection vaccines effective if given to individuals with nTB or aTB, we observed more pronounced differences between vaccines preventing any infectious disease and vaccines preventing any disease for current infection vaccines compared to any infection vaccines (Figure S 12, Table S 11).

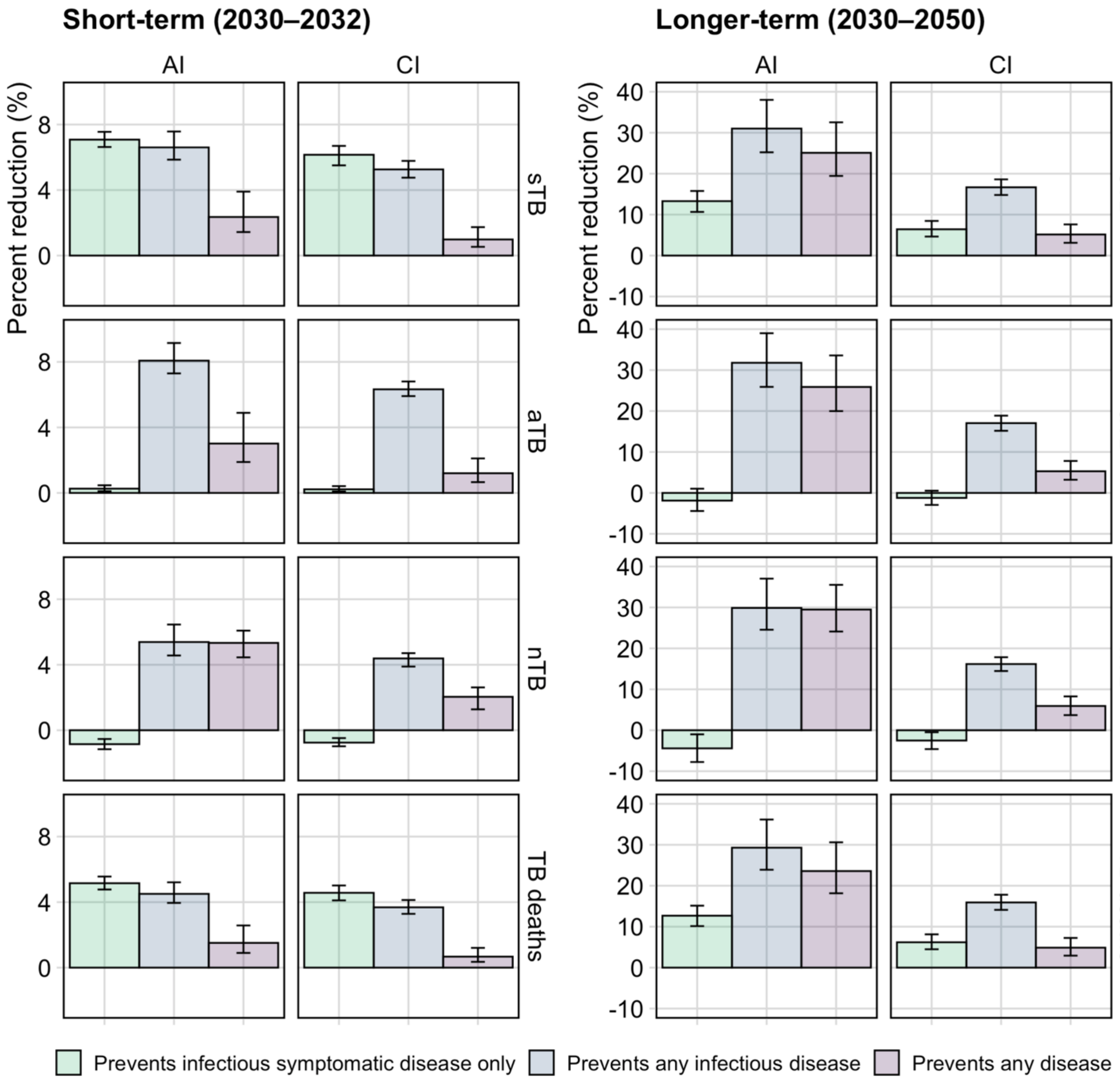


**Figure S 12 Percentage cumulative TB episodes and deaths averted between 2030–2032 (left column) or between 2030–2050 (right column), where vaccines were effective with any infection or current infection, including pre-symptomatic disease stages, with baseline (0.61–1) infectiousness of aTB relative to sTB.** Note y-axis scales differ. *Abbreviations: AI = any infection; aTB = infectious asymptomatic TB; CI = current infection; nTB = non-infectious TB; sTB = infectious symptomatic TB.*

**Table S 11 Number and percentage cumulative TB episodes and deaths averted (2030–2032 and 2030–2050) for vaccines effective in any infection or current infection, including disease stages, under baseline asymptomatic TB infectiousness relative to symptomatic TB.**

| | | AI (including disease) | | | CI (including disease) | | |
|---|---|---|---|---|---|---|---|
| Vaccine prevents progression to: | | Infectious symptomatic disease only | Any infectious disease | Any disease | Infectious symptomatic disease only | Any infectious disease | Any disease |
| Episodes Averted in millions (2030–2032) | sTB | 0.50 (0.44, 0.59) | 0.47 (0.39, 0.58) | 0.17 (0.10, 0.28) | 0.44 (0.38, 0.52) | 0.37 (0.32, 0.45) | 0.07 (0.04, 0.12) |
| | aTB | 0.07 (0.03, 0.12) | 2.15 (1.58, 2.74) | 0.80 (0.47, 1.40) | 0.06 (0.02, 0.11) | 1.69 (1.31, 2.08) | 0.32 (0.17, 0.56) |
| | aTB + sTB | 0.57 (0.48, 0.71) | 2.62 (1.98, 3.32) | 0.97 (0.57, 1.70) | 0.50 (0.41, 0.61) | 2.07 (1.64, 2.52) | 0.39 (0.21, 0.69) |
| | nTB | -0.30 (-0.37, -0.21) | 1.94 (1.32, 2.54) | 1.88 (1.27, 2.61) | -0.26 (-0.33, -0.18) | 1.56 (1.05, 2.11) | 0.70 (0.41, 1.10) |
| | TB deaths | 0.07 (0.06, 0.08) | 0.06 (0.05, 0.07) | 0.02 (0.01, 0.04) | 0.06 (0.05, 0.07) | 0.05 (0.04, 0.06) | 0.01 (0.00, 0.02) |
| % Reduction (2030–2032) | sTB | 7.08% (6.63, 7.55) | 6.60% (5.85, 7.57) | 2.36% (1.44, 3.90) | 6.16% (5.51, 6.70) | 5.26% (4.76, 5.78) | 0.98% (0.53, 1.73) |
| | aTB | 0.26% (0.10, 0.47) | 8.08% (7.30, 9.16) | 3.02% (1.89, 4.89) | 0.22% (0.09, 0.42) | 6.33% (5.91, 6.80) | 1.20% (0.66, 2.11) |
| | aTB + sTB | 1.69% (1.45, 2.02) | 7.76% (7.01, 8.82) | 2.88% (1.80, 4.69) | 1.47% (1.23, 1.78) | 6.10% (5.65, 6.59) | 1.16% (0.63, 2.03) |
| | nTB | -0.85% (-1.16, -0.53) | 5.39% (4.57, 6.46) | 5.33% (4.45, 6.08) | -0.75% (-0.98, -0.48) | 4.38% (3.89, 4.71) | 2.04% (1.28, 2.62) |
| | TB deaths | 5.15% (4.77, 5.56) | 4.50% (3.95, 5.21) | 1.51% (0.90, 2.57) | 4.57% (4.11, 5.01) | 3.68% (3.28, 4.13) | 0.67% (0.35, 1.20) |
| Episodes Averted in millions (2030–2050) | sTB | 6.22 (4.62, 8.32) | 14.63 (10.57, 20.14) | 11.81 (8.20, 16.61) | 3.02 (2.09, 4.13) | 7.86 (6.13, 9.88) | 2.43 (1.37, 3.85) |
| | aTB | -3.25 (-7.16, 1.90) | 55.86 (37.04, 79.25) | 45.26 (29.83, 66.27) | -2.19 (-4.64, 1.00) | 29.82 (21.72, 39.78) | 9.28 (5.25, 15.07) |
| | aTB + sTB | 2.88 (-2.00, 9.93) | 70.25 (48.53, 99.02) | 57.23 (38.36, 82.71) | 0.55 (0.37, 0.76) | 1.41 (1.09, 1.83) | 11.65 (6.65, 18.89) |
| | nTB | -10.41 (-16.67, -2.55) | 71.15 (46.49, 98.74) | 69.42 (44.66, 96.51) | -5.83 (-10.15, -1.21) | 37.28 (24.73, 54.85) | 13.91 (7.46, 22.53) |
| | TB deaths | 1.12 (0.83, 1.52) | 2.61 (1.93, 3.57) | 2.11 (1.47, 2.99) | 0.55 (0.37, 0.76) | 1.41 (1.09, 1.83) | 0.44 (0.24, 0.69) |
| % Reduction (2030–2050) | sTB | 13.29% (10.66, 15.77) | 31.02% (25.22, 38.03) | 25.09% (19.45, 32.54) | 6.42% (4.64, 8.46) | 16.67% (14.80, 18.61) | 5.15% (3.11, 7.61) |
| | aTB | -1.88% (-4.42, 1.02) | 31.78% (25.90, 39.01) | 25.88% (19.99, 33.58) | -1.22% (-2.93, 0.54) | 17.05% (15.17, 18.87) | 5.29% (3.22, 7.81) |
| | aTB + sTB | 1.29% (-0.92, 4.25) | 31.62% (25.77, 38.79) | 25.70% (19.87, 33.36) | 0.33% (-1.08, 2.23) | 16.97% (15.10, 18.81) | 5.25% (3.19, 7.76) |
| | nTB | -4.42% (-7.76, -1.00) | 29.89% (24.56, 37.03) | 29.48% (24.11, 35.52) | -2.51% (-4.60, -0.48) | 16.17% (14.47, 17.84) | 5.93% (3.70, 8.27) |
| | TB deaths | 12.69% (10.15, 15.13) | 29.30% (23.91, 36.17) | 23.58% (18.16, 30.61) | 6.19% (4.49, 8.13) | 15.92% (14.11, 17.80) | 4.87% (2.93, 7.24) |

*Abbreviations: nTB = non-infectious TB; aTB = infectious asymptomatic TB; sTB = infectious symptomatic TB*

### SUPPORTING DISCUSSION

This section should be read in conjunction with the main discussion.

We evaluated scenarios where we assumed, hypothetically, that the vaccines would be effective in pre-symptomatic disease stages at the time of vaccination. If this was the case, we saw greater impact from the vaccines due to protecting a larger proportion of the population, specifically those who were at a high risk of progressing to further disease stages. If aTB was assumed to have zero infectiousness, then the differences in impact between vaccine types on preventing sTB episodes was reduced. Over the longer-term, all vaccine types averted a similar proportion of sTB episodes, as accumulation of individuals in the aTB stage under the scenario where the vaccine only prevented progression to symptomatic disease would not result in continued transmission from aTB. Experts believe that it is unlikely that a vaccine would be effective if delivered to someone with disease at the time of vaccination, as the immune response would overwhelm any vaccine effect. However, whether the same applies for earlier disease stages (such as nTB) and undulation between disease stages is not yet known. Similarly, although limited in number, studies have suggested that aTB is likely to be infectious and may contribute to transmission (9,28).
